# From individuals to ancestries: towards attributing trait variation to haplotypes

**DOI:** 10.1101/2025.03.13.25323895

**Authors:** Yaoling Yang, Daniel J. Lawson

**Author notes:** Corresponding authors: Daniel J. Lawson and Yaoling Yang.

## Abstract

Genome-wide association studies (GWAS) have revolutionized our understanding of the genetic basis of complex traits and diseases, but limitations in SNP-centric approaches to population stratification limit the resolution of fine-scale population structures. Here we consider the use of haplotypes to represent population structure, leveraging haplotype components (HCs) for an improved understanding of trait associations and adjustment for population stratification. Using data from the UK Biobank, we showed that HCs have stronger associations with a range of phenotypes than principal components (PCs) while containing more predictive power for birthplaces globally. In GWAS, HCs-correction identifies more genome-wide significant association signals for birthplace- and lifestyle-related phenotypes, which are missed by PCs-corrected GWAS. Through thorough testing and simulation, we highlight challenges in performing ancestry-specific GWAS, underscoring the critical role of accurate local ancestry inference in studying admixed populations. We analyzed the haplotype structure of the UK Biobank in terms of 93 genetically-distinct populations, which enabled the computation of Ancestral Risk Scores (ARS) across 8 continental populations, providing insights into population-specific genetic risks for traits and diseases. By integrating haplotype information, this framework provides the potential to address challenges in population stratification, enhances GWAS resolution, and supports equitable health research by facilitating genetic studies in diverse populations.

## Introduction

GWAS have led to a great revolution in our understanding of the genetic basis of complex traits and diseases. These studies typically aim to identify associations between genetic variants and phenotypes, and they often require careful adjustment for population structure to avoid confounding. Population structure refers to the accumulated difference between ancestries due to independent evolution, which can bias GWAS results if it is not adequately adjusted^1^.

Whilst 40% of published associations in European ancestry are between SNPs and traits^2^, the SNP-centric perspective on population structure has several limitations in practice. SNPs change frequency only slowly, so careful modelling work is required to detect fine-scale structures^3,4,5^. Historically, principal component analysis (PCA) has been the most popular method for adjusting for population structure^6,7^. By accounting for population structure^8^, PCs help reduce confounding effects, therefore improving the accuracy of association estimates and minimizing false-positive findings.

In response to these limitations, other approaches have emerged that use the idea of haplotypes^3,9,10^ which leverage the inheritance process in which recombination breaks up the genome in long, continuous chunks - often measured as ‘identity by descent’^11^. Whilst for very recent timescales these describe family trees and pedigrees^12,13^, after just 20 generations (around 500 years) we each have 1 million ancestors, who can no longer be distinct, and the sharing of ancestors into the past behaves increasingly statistically. Learning such haplotype sharing allows the estimation of fine-scale population structure^3,4,5^.

A haplotype-centric view of population structure has many potential applications in understanding trait associations. This paper brings the discussion up to date and performs additional experiments to establish what haplotype approaches are likely to be useful, as well as discussing natural issues for which they may be less important. We will examine the informational content in the data, before considering uses in three broad categories: SNP associations via GWAS, ancestry of genomic segments, and whole-genome summaries based on ancestry-specific polygenic scores.

For correcting for population structure when learning genetic associations in GWAS, the most common approach is to correct for genome-wide genetic similarity, either measured through the Genetic Relatedness Matrix^14,15^ or PCA. These approaches all leverage SNP-level genetic similarity. Haplotype-centric approaches have been recently developed, with ancestry components^16^ comparing a GWAS dataset to a pre-defined set of reference populations to identify haplotypes. Haplotype-sharing patterns learned within the GWAS dataset measured by HCs^17^ are directly comparable to PCs. By leveraging the computational efficiency of the ‘Positional Burrows-Wheeler Transform’^18^, HCs become well-suited for large datasets such as the UK Biobank^17^. Notably, HCs predict top PCs and retain information that is not captured by PCs, such as finer-scale associations with birthplaces, as Fig. 1 illustrates (discussed completely in Results).

**Fig. 1:**
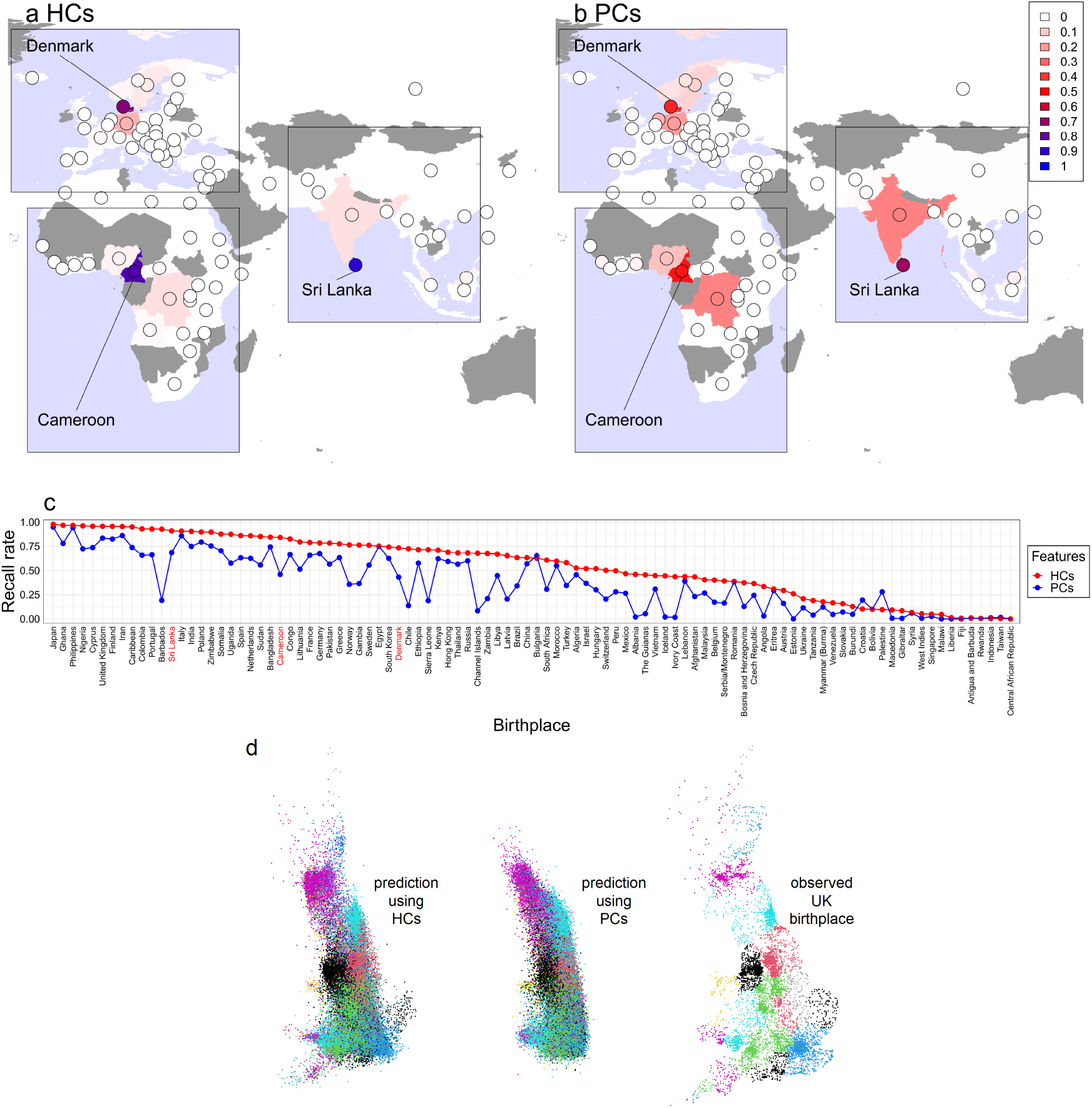
Comparison of the performance of HCs and PCs for predicting birthplace in a left-out sample. a-b, the prediction of Denmark, Cameroon and Sri Lanka by HCs and PCs visualized in a world map. c, the recall rate of each birthplace as predicted by HCs and PCs analyzing n=20,092 internationally born participants from the UK Biobank with non-British ancestry. d, visualization considering n=341,881 British-born individuals with self-reported British ethnicity, coloured by ancestral region of birthplace.

Building on these initial findings, our first goal is to systematically compare the efficacy of HCs and PCs in representing genome-wide ancestry and controlling for ancestral confounders in GWAS and related techniques. We assess predictive power across a wide range of continuous and binary phenotypes in the UK Biobank^19^, using regression models to compute the coefficient of determination (*R*^2^) for each approach. In addition, expanding on the success of birthplace prediction within the UK^17^, we broaden this comparison to include worldwide populations. We also perform GWAS on UK Biobank traits correcting for population stratification using each measure to identify SNPs that show significant differences in association when controlled with HCs versus PCs.

The second goal is to consider applications in admixed datasets. Whilst migration and globalization lead to more people having ancestry from multiple ancestral populations, these admixed individuals have often been excluded from large-scale genomic studies due to challenges related to population structure^20^. The complexity arises because population substructure can lead to false positives in GWAS if not effectively controlled. Conventional approaches, like using PCs to adjust for broad population structure, are ineffective for admixed populations because they only capture global ancestry proportions and not the specific local ancestry at each genomic locus^21^. As a result, the inability to handle local ancestry has limited the clinical utility of large-scale genomic datasets for admixed populations, contributing to ongoing health disparities^22,23,24^.

Local ancestry inference (LAI)^17,25,26,27,28^offers hope for working with admixed populations. Several attempts have been made to leverage LAI to examine the trait-gene associations, such as local ancestry PC correction^29^, the inclusion of local ancestries as covariates to reduce confounding^30^, and a two-step local ancestry adjusted testing procedure named LAAA^31^. Methods such as AsaMap do not rely on accurate LAI^32^ but do not considerably improve power compared with standard GWAS.

More recently, the Tractor framework^21^ enables the inclusion of admixed individuals in GWAS by accounting for both the global and local ancestral composition of each individual, providing ancestry-specific effect-size estimates. Since we can now compute LAI at scale it is natural to deploy Tractor-like models on biobanks. By simulating different levels of uncertainty in local ancestry calls, we aim to understand how these uncertainties affect the performance and accuracy of the Tractor model in estimating ancestry-specific effect sizes. These simulations are crucial for assessing the feasibility of using Tractor in practice, but unfortunately, we will show that uncertainty in LAI prevents estimating ancestry-specific effect sizes for rare SNPs in this framework. Fortunately for our understanding of the underlying biology, there is increasing evidence that the majority of traits lack ancestry-specific effects^16^.

Our final goal is to use local ancestry to estimate ancestral contributions to genetic traits. The comparison of homogeneous populations with different ethnicities has revealed different genome-wide trait associations or genetic scores^33,34^. Given access to local ancestry information, it is natural to attribute trait association across ancestries in an ‘ancestral risk score’^35,36^. We will show that ARS can quantify the genetic risk of each ancestry accounting for local ancestry probabilities. This flexibility makes ARS particularly suitable for studies with high uncertainty in local ancestry estimates, as it can still provide meaningful insights into ancestry-specific risks without requiring definitive ancestry calls at each locus. In this work, we painted the UK Biobank using a fine-scale reference panel with 4,334 reference individuals from public-available data sources (i.e. all except POPRES) as used in Hu et al.^16^, and applied ARS to compute ancestry-specific risk scores across multiple diseases.

## Methods

### Prediction of worldwide birthplaces in the UK Biobank with HCs or PCs

We aimed to assess the ability of HCs and PCs to predict individuals’ birthplaces. We summarized the number of participants in the UK Biobank from different worldwide birthplaces by their self-reported ethnic group (hereafter ‘ethnicity’), i.e. British, white (excluding British), Asian, black, mixed, others or unknown. For each birthplace, we included participants from ethnicities representative of local residents only if there were at least 10 individuals for those ethnicities. Additionally, we sampled 1,000 individuals born in the UK to balance the dataset. These filtering steps resulted in a final cohort of 20,092 individuals (including both males and females) from 98 birthplaces, which were summarized in Supplementary Table 1.

We conducted an analysis to compare the prediction performance of the top 150 HCs and PCs on birthplaces with eXtreme Gradient Boosting (XGBoost^37^). First, we randomly split the datasets with stratified sampling into an 80% training set and a 20% test set. We performed a 5-fold cross-validation (CV) on the training data to determine the optimal maximal number of boosting iterations, where 4 folds were used for training and 1 fold for validation. Then, XGBoost models were trained using 150 HCs or PCs and categorical birthplace labels in the training set, and predictions were made on the test set. This process was repeated 50 times, after which the average confusion matrix was computed for further analysis, including the visualization of the prediction results in a heatmap (Supplementary Fig. 1) or a world map (Fig. 1a-b).

We repeated this analysis for the 341,881 individuals who were born in the UK and self-identified as white or British ethnicity, applying XGBoost to the Easting and Northing components to predict UK birth locations, and reported the visualization results in Fig. 1d.

### Computation of loadings and autocorrelation of HCs and PCs

Let *X* denote the *N* × *L* genotype data for *N* = 406, 773 unrelated individuals (including both males and females) within the UK Biobank, as implemented by Bycroft et al.^19^, and *L* = 147, 604 SNPs. Let *H* denote the *N* × *K* HC matrix for the top 150 HCs (following the computation of Yang et al.^17^). We aimed to project *H* onto *X* to compute the SNP loadings 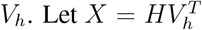 be the decomposition of *X* in terms of HCs. We computed *V*_*h*_ = ((*H*^*T*^ *H*)^−1^*H*^*T*^ *X*)^*T*^ by matrix transformation.

To compute the loadings for PCs *V*_*p*_, we performed the singular value decomposition on *X*, i.e. 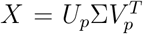 where Σ is a diagonal matrix of the singular values, and then we extracted *V*_*p*_ with the top 150 components, i.e. PCs.

Next, we estimated the autocorrelation function (ACF) for both the absolute value of loadings for HCs *V*_*h*_ (after normalization to ensure it has the same scale as *V*_*p*_) and PCs *V*_*p*_, and visualized the average ACF at lag 1 in Supplementary Fig. 2a. We also visualized the absolute value of loadings for specific HCs and PCs throughout the genome with SNPs aggregated into bins of 100 (Supplementary Fig. 2b).

**Fig. 2:**
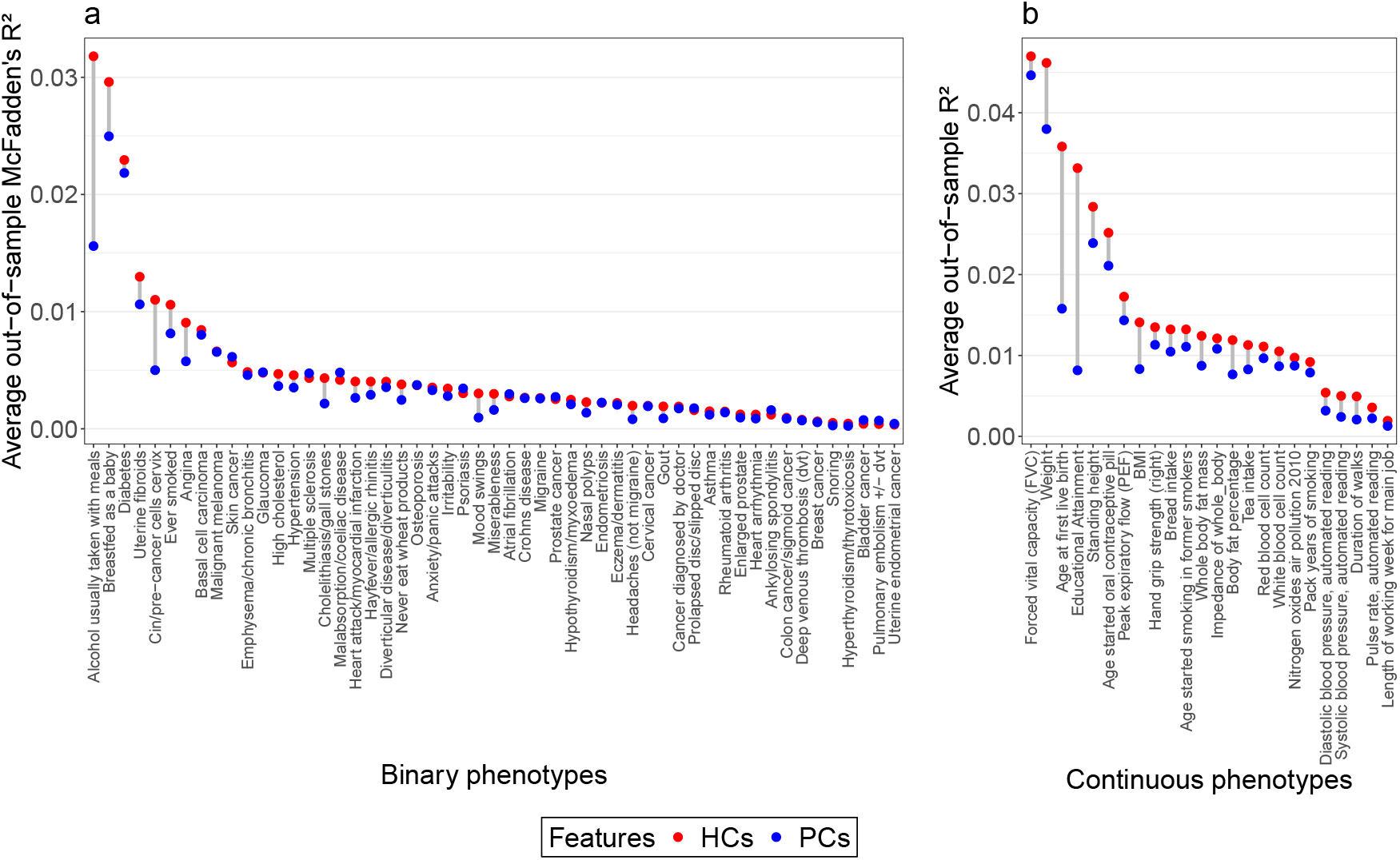
Comparison of the predictive performance of HCs and PCs on phenotypes. We visualize the average out-of-sample *R*^2^ explained by HCs (red) and PCs (blue) under linear regression models (plot a, for continuous phenotypes) or logistic regression models (plot b, for binary phenotypes) through a 5-fold CV performed on n=462,694 individuals in the UK Biobank, constraining to PCs not associated with genomic structure.

### Comparison of prediction performance between HCs and PCs through cross-validation

To evaluate the information contained in HCs/PCs data, we compared the predictive performance of the top 150 HCs and PCs by fitting regression models on 24 continuous and 53 binary phenotypes. For each model, we used either PCs or HCs as predictors and included sex and age as covariates. Instead of simply reporting the in-sample *R*^2^, we performed a 5-fold CV scheme and reported the average out-of-sample *R*^2^ on the test set. In detail, we trained the model using 4 folds and tested it with 1 fold, and then we repeated the process 5 times until each fold was used as the test fold once.

We also aimed to find the optimal number of HCs/PCs with the best out-of-sample performance. Here only the top 18 PCs were considered because the remaining PCs capture complex LD structure rather than solely population structure (Supplementary Fig. 2)^16,8^. In specific, we applied the above CV process for any of the top k HCs/PCs (k=1,..,150 for HCs and k=1,..,18 for PCs), and reported the optimal k with the highest average out-of-sample *R*^2^.

Different measurements of *R*^2^ were used for different models. We performed linear regression models on continuous phenotypes and reported the standard *R*^2^; for binary phenotypes, we performed logistic regression models and used McFadden’s pseudo-*R*^2^ to measure the variance explained by the model. To measure the *R*^2^ of only the HCs and PC while removing the effects of sex and age, we used the subtraction of the *R*^2^ of the full model (i.e. *Y* ∼ sex + age + HCs*/*PCs) and the *R*^2^ of the model with only sex and age as covariates (i.e. *Y* ∼ sex + age).

The study was performed on 406,773 unrelated individuals (including both males and females) within the UK Biobank, as implemented by Bycroft et al.^19^

### GWAS scan with HCs or PCs correction

To compare the effectiveness of HCs and PCs in correcting for population structure in GWAS, we conducted GWAS scans on the aforementioned 24 continuous and 53 binary phenotypes, plus the east and north coordinates of the birthplace, using PLINK 2.0^38^. The sex and age of participants are also included in the GWAS model as the standard confounding correction. The top 18 HCs and PCs are used for genome-wide population correction, respectively. Standard linear regression models are used for continuous phenotypes, and logistic regression models for binary phenotypes.

### Public-available reference panel with 93 populations

We summarized the public-available reference panel following the steps in Hu et al.^16^, including HAPMAP3^39^, HGDP^40^, POBI^5^, Busby Europe^41^, Pagani Africa^42^, Peterson Africa^43^, Schlebusch - Africa^44^, and excluding POPRES^45^ (due to restricted access), which contains 4,334 individuals (including both males and females) from 129 worldwide populations. Below we performed an additional quality control on this reference panel.

We applied PBWTpaint to compute the expected length of genome matched (or ‘copied’), which approximates the amount of genome the *i*th individual shares most recently with the *j*th, hereafter ‘chunk length’, denoted *A*_*ij*_. This process requires phased genotype data. In detail, for each haplotype, we then calculated the sum of the chunk lengths weighted by the total genetic distance of each chromosome. This was performed for both haplotypes of each individual, after which we combined the two haplotypes to obtain a total measure per individual. We use *A*_*ij*_ to denote the total genomic length that the *i*th individual copied from the *j*th individual.

Using the population labels of each individual provided by Hu et al.^16^, we aggregated *A*_*ij*_ values by population, and therefore *A* shrinks from an *N* × *N* matrix to a *K* × *K* matrix, where *N* is the number of individuals and *K* is the number of populations. Next, we normalized each row by the diagonal element which refers to the expected length of copied chunks from the same population. Because we have fewer samples than Hu et al.^16^, not all populations are as cleanly separated. We therefore merged populations for which the inter-individual chunk length variation overlapped iteratively until each updated population was well distinguished from others. This resulted in a refined reference panel with 93 distinct populations (Supplementary Table 2).

### Paint the UK Biobank with the public-available reference panel

We inferred the local ancestry of UK Biobank individuals using the public-available reference panel as the reference data, which includes 4,334 individuals from 93 populations. We filtered the common bi-allelic SNPs with minor allele frequency MAF ≥ 5% in both the imputed UK Biobank dataset and the reference dataset, resulting in a total of 667,543 SNPs. Next, these two datasets were merged and then phased using Beagle 5.4^46^. Finally, we split the phased dataset into the reference and target datasets according to their labels.

We did an additional filtering of the UK Biobank individuals by removing individuals who have at least one of the grandparents in common. In detail, we computed the pairwise genetic relatedness score within each chromosome using the ‘–genome’ command with PLINK 2.0^38^, and then computed the average of them weighted by the genetic distance of each chromosome, which was denoted as the genome-wide pairwise genetic relatedness score *S*_*ij*_. We removed the least number of individuals to ensure that *S*_*ij*_ between any individual *i* and *j* is smaller than 0.24 (i.e. relative removal excluding cousins and grandparents, whilst retaining some rare populations with high inbreeding coefficients). The final UK Biobank dataset has 462,694 individuals (including both males and females).

The local ancestry inference implemented by SparsePainter^17^ requires the input of parameter ‘fixlambda’, i.e. the recombination scaling constant. We estimated it as 128.6 using the Viterbi algorithm on chromosome 20 and used this fixed value for the LAI across all the chromosomes. We set the parameters of SparsePainter to find the 20 longest matches (longer than 10 SNPs) at each SNP, and we used the default setting for all the other parameters. SparsePainter paints in batches which we set to 10,000 individuals to trade-off parallelism with memory storage and data input.

### Simulation for ancestry-specific GWAS

With local ancestry inferred, we are interested in estimating the ancestry-specific effect sizes for GWAS. With accurate and certain local ancestry calls, Tractor^21^ has been shown to accurately estimate ancestry-specific effect sizes. At a given SNP, let *Y* denote the phenotype, *T*_*k*_ denote the number of copies of the risk allele from ancestry *k*, and *X*_*k*_ denote the number of copies at this locus (i.e. either the risk allele or the alternative allele) from ancestry *k*. Also let *C*_*j*_(*j* = 1, 2, ..*q*) denote *q* different covariates, and *ϵ* denote the random error. Under a two-way admixture, the Tractor model for continuous outcome is:

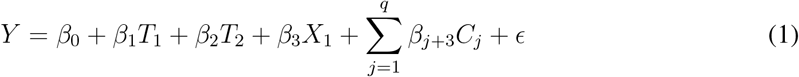

However, in practice, local ancestry is not known and must be inferred. Many modern local ancestry inference tools, such as SparsePainter^17^, ChromoPainter^3^, RFMix^27^ and FLARE^25^ implement Li and Stephens hidden Markov model^47^ and therefore can report local ancestry probabilities at each locus. Other software, including FLARE, report ancestry calls as a best guess of local ancestry probabilities, omitting loci that were admixed longer ago and hence are uncertain. Therefore, we performed a simulation study to compare the performance of the Tractor model under different LAI uncertainties, with *T*_1_, *T*_2_ and *X*_1_ computed from either (a) The true local ancestry call (hereafter ‘TRUTH’); (b) The raw local ancestry probabilities (hereafter ‘RAW’); (c) The best-guess local ancestry call from raw local ancestry probabilities (hereafter ‘BESTGUESS’); or (d) Random sampling of local ancestry from raw local ancestry probabilities (hereafter ‘SAMPLING’).

Below we describe our simulation procedures. The sample size for the simulation is *N* = 20, 000 diploid individuals. For simplicity, we simulated two ancestries and therefore considered only the expected value of Ancestry 1 that *a* = {0.1, 0.25, 0.5}. To generate individual variation in certainty with a controlled mean that can be interpreted as an ‘error rate’, we first sampled a latent variable *Ã*_*ikd*_ for the *d*th copy of the *i*th individual for the *k*th ancestry, from Bernoulli distribution: *Ã*_*i*1*d*_ ∼ Bernoulli(*a*) and *Ã*_*i*2*d*_ = 1 − *Ã*_*i*1*d*_. Since the haplotypes are not observed perfectly, we considered different average confidence, i.e. the probability of LAI: *p* = {0.5, 0.6, 0.7, 0.8, 0.9, 0.95, 0.98} which are assumed to be correctly calibrated. We sampled the haplotype certainties *p*_*id*_ for the *d*th copy of individual *i* from Beta distribution with mean *E*(*p*_*id*_) = *p* and variance *V ar*(*p*_*id*_) = (0.05*/p*)^2^. This forms the ‘observed’ local ancestry probability *p*_*ikd*_ = *p*_*id*_ if *Ã*_*ikd*_ = 1, and *p*_*ikd*_ = 1 − *p*_*id*_ otherwise. Finally, we sampled the true ancestry call 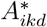 from Bernoulli distribution: 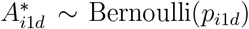 and 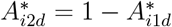.

We simulated the genotype *G*_*id*_ for the *d*th copy of individual *i* using the Balding-Nichols model^48^ with a fixation index of *F*_*st*_ = 0.2, which accounts for varying local ancestry *A*_*ikd*_. We considered SNPs falling into 4 different MAF thresholds: 1% ≤ *f* ≤ 5%, 5% ≤ *f* ≤ 10%, 10% ≤ *f* ≤ 20%, and *f* ≥ 20%. The ancestral frequency *f* is Uniform under each MAF threshold, from which the population-specific risk allele frequency for the *k*th ancestry was simulated as 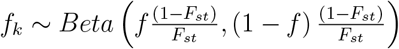 also with restrictions of the corresponding MAF threshold. The alleles *G*_*id*_ were simulated from Bernoulli distribution with their population’s frequency: *G*_*id*_ ∼ Bernoulli(*f*_*k*_), where *k* = 1 if 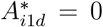,and *k* = 2 if 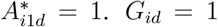 and *G*_*id*_ = 0 represent the risk and alternative allele, respectively.

Subsequently, we simulated a trait from the Tractor model using the true haplotype ancestries, i.e. 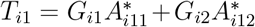 and 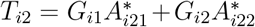 as the true ancestral dosages, and then generated the phenotype *Y*_*i*_ = 2 + 5*T*_*i*1_ + *T*_*i*2_ + *ϵ*_*i*_, where *ϵ*_*i*_ ∼ *N* (0, 15^2^). We used a high variance of the random error because the heritability of a single SNP in practice is always very low.

We then constructed the genetic effect values under the same model but using the *observed* ancestry under different treatments of uncertainty, i.e. 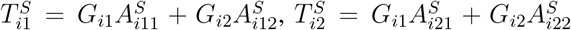 and 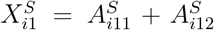, where *S* = {TRUTH, RAW, BESTGUESS, SAMPLING}. Intuitively, 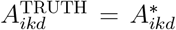, and 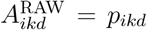 .For BESTGUESS, we let 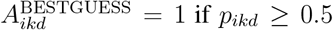, and 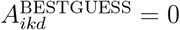 otherwise. For SAMPLING, we sampled from Bernoulli distribution : 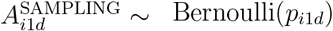 and 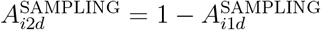.

Finally, we fit the linear regression model 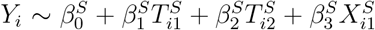.For each scenario, we simulated 1,000 traits and compared the average estimated 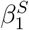 and 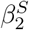 with the truth, i.e. *β*_1_ = 5 and *β*_2_ = 1.

### Computing the distribution of haplotypes across ancestral probabilities by continent

For the subsequent analysis, we focused on 35 phenotypes (the majority of them are diseases) which were reported (in the UK Biobank database) to have at least 30 SNPs significant at *p <* 5 × 10^−6^ that overlap our SNP set before and at least 5 after linkage disequilibrium (LD) pruning. We painted the UK Biobank using the pre-described reference panel, and summarized the painting from 93-population level into 8-continental level, i.e. Europe (all Europe excluding East Europe and UK), East Europe, UK, Africa, South Asia, Middle East, East Asia, and America (see Supplementary Table 2 for detail). Then we aimed to assess the distribution of haplotypes across ancestral probabilities by continent. For each phenotype, we selected the most significant SNP, and haplotype counts (with a total of 925,388, i.e. twice the number of individuals) exceeding specific ancestral probability thresholds ranging from to 1.0 (with a step size of 0.01) were computed for all continents (Supplementary Fig. 3). Median counts and 95% confidence intervals were calculated to summarize the distribution of haplotype numbers at each threshold.

**Fig. 3:**
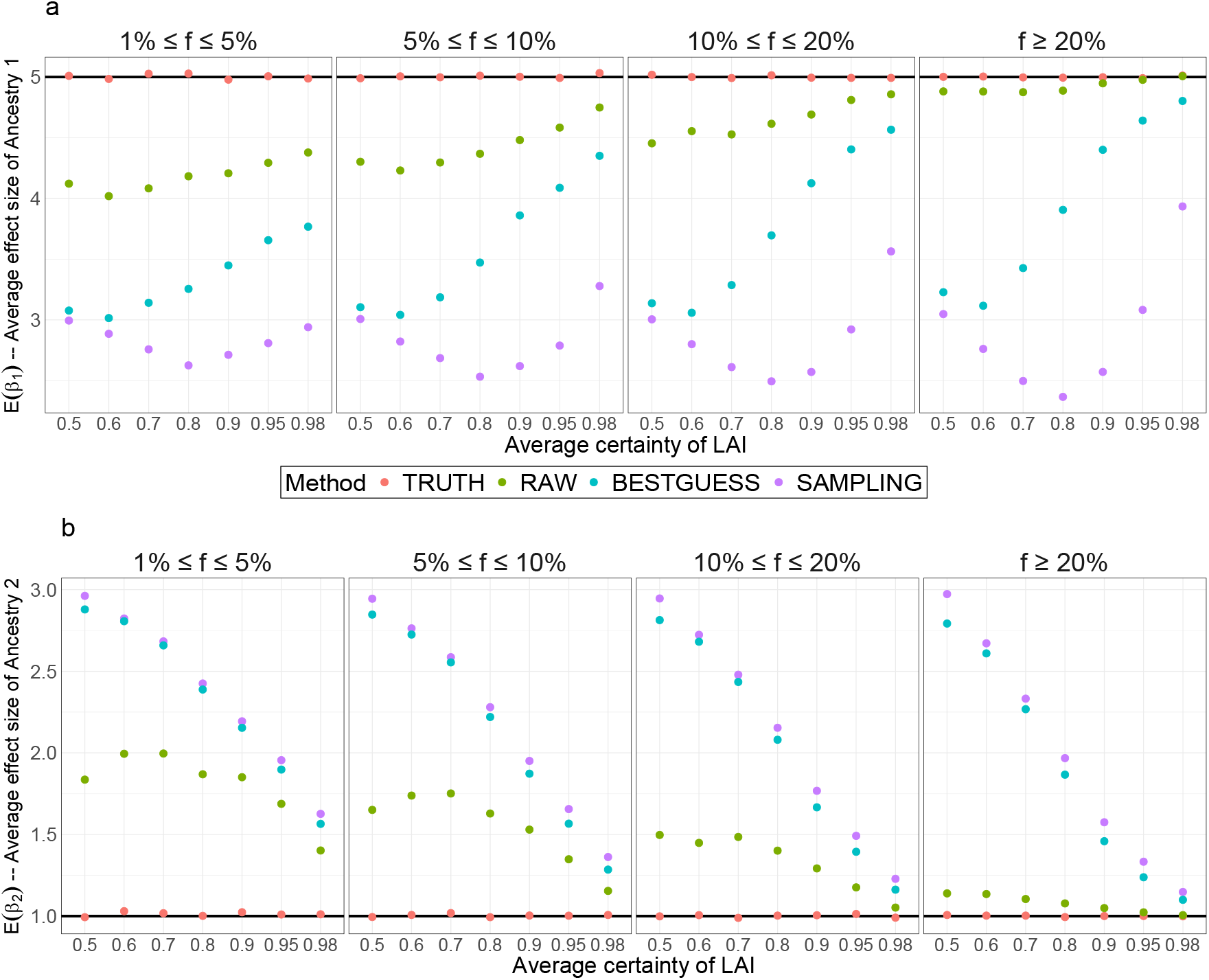
Comparison of the average estimated effect size of ancestries from the Tractor model with different methods to represent local ancestries under. 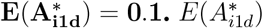 denotes the average probability of Ancestry 1. The x-axis represents the average certainty of LAI (see Methods) and the y-axis represents the average estimated effect sizes for Ancestry 1 (plot a) and Ancestry 2 (plot b). Different MAF thresholds *f* are compared: 1% ≤ *f* ≤ 5%, 5% ≤ *f* ≤ 10%, 10% ≤ *f* ≤ 20%, and *f* ≥ 20%. The simulation was repeated 1,000 times with n=20,000 diploid individuals.

### Computation of ARS

We computed the ARS^35,36^ for 35 phenotypes at the 8 continental levels described above. We first filtered all the SNPs which are significant at *p <* 10^−6^ from the published GWAS results and are contained in our SNP set, and then did LD pruning in PLINK 2.0^38^ based on *R*^2^ at a threshold of 0.5. Let *A*_*ijk*_ denote the local ancestry probability of ancestry *k* for the *i*th haplotype at the *j*th SNP, and *G*_*ij*_ denote the genotype of the *i*th haplotype at the *j*th SNP, which takes value 1 or 0 representing the risk or the alternative allele, respectively. Then the risk allele frequency of SNP *j* for ancestry *k, f*_*jk*_, is computed as:

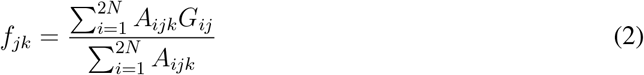

where *N* is the total number of individuals.

We fit linear and logistic regression models for continuous and binary phenotypes, using sex, age and the top 150 HCs as covariates, and computed the effect size *β*_*j*_ for the risk allele of the *j*th SNP. To compute ARS for ancestry *k*, we summed over all *M* pruned significant SNPs in an additive model:

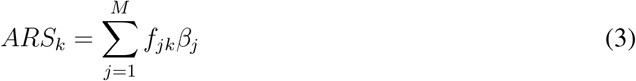

### Uncertainty for ARS

There are two types of randomness involved in the calculation of the ARS. The first is ‘randomness over which individuals were in our dataset’. Calculating ARS involves estimating the frequency of a trait-associated SNP in each of the populations, some of which have few samples. The second is ‘randomness over the ancestry assigned to trait-associated SNPs’. As relatively few SNPs are sampled for some traits, we might expect a similar random set of SNPs to contain spurious trait associations.

#### Randomness in ARS over individuals

To obtain a 95% confidence interval for an ARS accounting for the random sample of individuals, we bootstrapped over individuals for 1,000 times. We obtained the 2.5% and 97.5% quantile of risk allele frequency of SNP *j* for ancestry 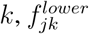 and 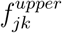,which replace *f*_*jk*_ in Equation (3) to obtain the lower and upper bound of ARS for ancestry *k*, 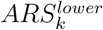 and *A*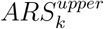.

#### Randomness in ARS-ancestry associations

To account for variation in ancestry assigned to trait-associated SNPs, we simulate pseudo-ARS under the null that trait-associated SNPs are not associated with ancestry. The pseudo-ARS are generated by matching each associated SNP with a set of LD-pruned SNPs with the same risk allele frequency (precision up to 1%) in the UK Biobank.

To illustrate the variation in SNP ancestries genome-wide, we visually compare the value of the ancestry of each SNP with its pseudo-replicates. Supplementary Fig. 4 shows this per-SNP for the 8 pruned significant SNPs for pulmonary embolism. Some visually extreme ancestral frequency estimates are expected under the null, whilst many significant SNPs for pulmonary embolism are outliers of allele frequencies in specific populations. Whilst this large *F*_*st*_ may indicate selection, it only translates to a signal for the target trait if it replicates at other SNPs.

**Fig. 4:**
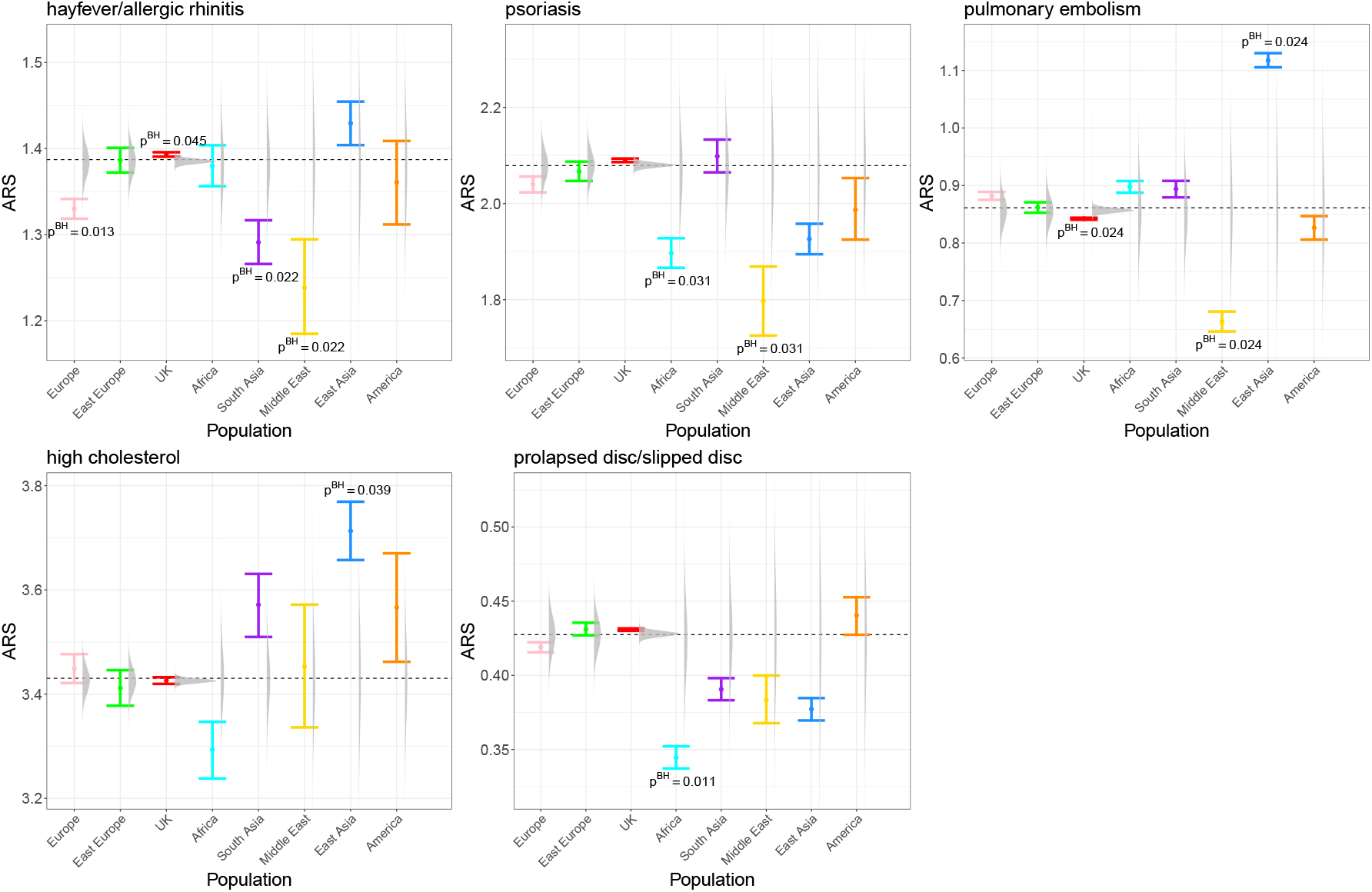
ARS for 5 significant phenotypes ordered by the overall significance level. ARS are computed from n=462,694 individuals in the UK Biobank. The error bars represent the 95% confidence interval. The distribution of simulated ARS for each population is shown as a raincloud plot, under the null hypothesis that SNPs associated with the trait are not associated with ancestries (Methods). The dashed black lines represent the average ARS of all populations weighted by the population sizes. The ARS of populations with Benjamini-Hochberg corrected p-values significant at 5% computed from the two-sided empirical test are annotated.

### Assessing the significance level of ARS

To assess whether specific ARS are significant, we combine the above two causes of variation. For each pruned significant SNP *j* of each phenotype, we found all the SNPs across all 22 chromosomes with the same allele frequency (precision up to 1%), and used LD pruning to remove chance duplications. Then we bootstrapped over SNPs, i.e. resampled with replacement, for 50,000 times, and obtained the simulated risk allele frequency 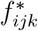 for the *k*th population, where *i* = 1, …, 50000 denotes the index of the bootstrapped SNPs. Next, we used the real risk-allele effect size *β*_*j*_ to compute the simulated 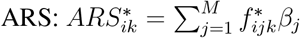.

We then designed a test for whether ARS for the *k*th population is significantly different from the population mean under the null that the variation observed is driven by uncertainty in frequency estimates (bootstrap over individuals) and randomness in which SNPs are associated with ancestry. For this, we sampled a realization of the ARS accounting for uncertainty: 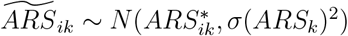,where *σ*(*ARS*_*k*_) is the standard error obtained by bootstrap over individuals. From this, we can compute an empirical test statistic accounting for randomness in SNP ancestry calls. We computed a two-sided p-value for each population as two times the proportion that (observed, individual-variation accounted) 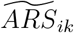 is more extreme than (ancestry sampling of SNP accounted) *ARS*_*k*_ (Supplementary Fig. 5). We then used Benjamini-Hochberg correction^49^ on the p-values to control the false discovery rate.

We finally used Fisher’s method to combine the p-values *p*_*k*_ of all *K* = 8 populations as an aggregated p-value for each phenotype. In detail, we computed the test statistic 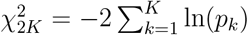, and then computed the p-value for 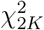 with 2*K* degrees of freedom. Finally, the Benjamini-Hochberg corrected p-values are reported in Supplementary Fig. 6.

## Results

### Prediction of worldwide birthplaces using HCs and PCs

To illustrate the population structure captured by HCs and PCs, we assessed their ability to predict individuals’ birthplaces using a dataset of 20,092 non-British ethnicity UK Biobank participants from 98 worldwide birthplaces (Methods). Using XGBoost models trained on the top 150 HCs or PCs, we found that models based on HCs had a notably higher average recall rate (80.19%) than those based on PCs (62.68%), indicating that HCs have stronger associations with birthplaces than PCs. The average probabilities of correctly predicting each birthplace, i.e. the recall rate, were visualized in Fig. 1c and confusion matrices in Supplementary Fig. 1, which highlights the superior predictive performance of HCs in capturing fine-scale population structure. For example, PCs failed to distinguish Denmark from nearby European countries, and Cameroon from adjacent African countries, and PCs mistake a great proportion of Sri Lanka people as Indian (Fig. 1b). By contrast, HCs show much higher prediction accuracy in those birthplaces (Fig. 1a). Fig. 1d applies the XGBoost pipeline within the UK to visualize how HCs capture subtle population structure lost by PCs, including drift components from Eastern, South Eastern, and South Western England and North-East Scotland.

### Comparison of UKB phenotype prediction performance of population structure represented by HCs and PCs

HCs have at least two advantages over PCs. The first is conceptual - because PCs are constructed from total variance explained, many are associated with genetically localized structures rather than population structure. HCs, being built on recent relatedness patterns, are immune to this and so any number can be used in principle. To illustrate this, Supplementary Fig. 2 shows how every PC¿18 is localized to one or a few genomic regions as summarized with autocorrelation, with e.g. PC19 tagging a single region on chromosome 6. The second is practical - as they represent more recent relationships in finer-scale, they can be expected to capture more population structure.

To assess the information content in HCs/PCs (chosen by cross-validation), we compared the performance of using up to 18 PCs (these first 18 describe genome-wide average SNP frequency variation, whilst the rest are also mixed with genomic structure, as shown in Supplementary Fig. 2 and ref.^8,16^) and 150 HCs (which describe within-sample recent relationships via haplotypes) to predict phenotypes in left-out data. In out-of-sample performance, HCs explain higher variance than PCs for all continuous and nearly all binary phenotypes (Fig. 2, Supplementary Tables 3-4), for instance, 4 times for ‘educational attainment’, over twice for ‘age at first live birth’, ‘alcohol usually taken with meals’, and ‘cin/pre-cancer cell cervix’. Other notable increases are ‘breastfed as a baby’, ‘weight’, ‘BMI’, ‘angina’, ‘cholelithiasis/gall stones’, ‘mood swings’, etc.

To test whether constraining PCs mattered, we computed the out-of-sample performance without constraint, for top HCs/PCs for every phenotype (Supplementary Figs. 7-8), with largely unchanged qualitative conclusions. No continuous phenotype could be as well explained with PCs as with HCs, and for those few binary traits where HCs are similar or marginally outperformed by PCs, the total variance explained is tiny because those traits are very rare. As British populations dominate the UKB, the significant increase of *R*^2^ observed for certain HCs suggests that these components capture variations specific to the UK. By contrast, many other HCs associated with under-represented populations in the UKB may simply add noise when predicting phenotypes.

By capturing pairwise coancestry, HCs can offer a finer-scale representation of population structure (and associated geographical or socio-economic confounding) than traditional PCs, thus leading to better variance explanation ability in GWAS and related analyses.

### GWAS with HC and PC correction

To compare the effectiveness of HCs and PCs in correcting for population structure in GWAS, we performed GWAS on 26 continuous and 53 binary phenotypes, and visualized them in Supplementary Figs. 9-10.

For educational attainment, we found 6 SNPs overlapping gene ACMSD and CCNT2-AS1 which were significant at genome-wide significance level with HC-corrected GWAS, but had no significance (*p >* 0.01) with PC-corrected GWAS. Through checking the UK Biobank database, these SNPs were also reported to have GWAS hits in many other phenotypes, which indicates that these might be real signals over-corrected by PCs. Because HCs are highly informative of birthplaces, we found changes in significance when correcting for HCs/PCs in both the east and north coordinates in a number of SNPs.

However, in the UK Biobank dataset, no substantial differences were observed between HC- and PC-corrected GWAS for other phenotypes, indicating that HCs have advantages primarily in specific contexts where fine-scale population structure is important. This may include datasets that include inter-continental admixture, which is of limited scale in these data.

### Simulation reveals ancestry-specific GWAS requires accurate local ancestry assignment

We intended to estimate ancestry-specific effect sizes in the UK Biobank. To evaluate whether these results would be meaningful, we therefore performed simulations to evaluate methods for estimating ancestry-specific effect sizes in admixed populations. Using a two-way admixture model with varying ancestry proportions, we compared the performance of the Tractor model^21^ with different MAF thresholds and LAI certainties under different methods of representing local ancestry: TRUTH, RAW, BESTGUESS, and SAMPLING (see Methods).

The Tractor framework can always return accurate estimates of effect sizes on average (Fig. 3 and Supplementary Figs. 11-12) when we know the true ancestries (TRUTH), while SAMPLING and BESTGUESS are always inaccurate. By contrast, although replacing definitive ancestry calls in the Tractor framework with the local ancestry probabilities (RAW) achieves high accuracy under high certainty (≥ 0.9) when modelling common SNPs, i.e. MAF ≥ 20%, the estimates deviate considerably from the truth under imperfect LAI for less common SNPs. We further revealed that few haplotypes outside of Europe meet the threshold of 95% and 98% LAI probability (Methods, Supplementary Fig. 3), indicating the limited power of the UK Biobank to implement the Tractor framework in practice effectively.

Moreover, the RAW approach has substantially higher standard errors in effect size estimates (Supplementary Figs. 13-15), which consequently reduces the statistical significance in practical GWAS. Given that significant GWAS signals often arise from rare SNPs, these findings indicate the challenges of accurately estimating ancestry-specific effect sizes with the Tractor framework under conditions of LAI uncertainty.

We have also explored using Bayesian modelling or variational inference to estimate the ancestry-specific effect sizes. However, with 2*N* (*k* − 1) ancestry calls to estimate, which exceed the sample size *N*, accurately inferring the ancestry-specific effect sizes becomes almost infeasible. Real GWAS are heterogeneous and causal loci cannot be assumed to be the high-frequency tagging variants that have the most predictive power. Therefore, we conclude that while Tractor serves as an idealized model, its utility is primarily limited to contexts involving highly distinct populations. In fine-scale admixed populations, its effectiveness depends heavily on the precision of local ancestry assignments, which remains a challenge with current tools. Thus, Tractor may still be applicable in admixed populations, provided the ancestral differences are clear enough and can be accurately identified - which we will show is not true of the UK Biobank.

### ARS scan for traits across continental populations detect ancestral-specific risks

To work with quantities that are uncertainty-tolerant, we painted the UK Biobank with a public-available reference panel with 93 populations that we summarized (Methods, Supplementary Table 2), and then computed ARS for phenotypes across 8 continental populations to assess population-specific genetic risk profiles, and quantified their uncertainty.

We computed p-values using a two-sided empirical test (Methods) that accounts for variation in individuals and SNPs. We tested the null hypothesis that our associations are driven by chance sampling of ancestries for associated SNPs, both for single SNP associations (Supplementary Fig. 4) or to identify ARS signals that are real outliers (Fig. 4). Visualization of these results shows variability in the ARS estimate for the SNPs actually associated with the trait (confidence intervals) as well as ‘simulated ARS’ that could be expected by matching SNPs with equal frequency genome-wide (grey raincloud plots). The test accounts for both of these effects together.

To assess the overall significance of each phenotype, we employed Fisher’s method and adjusted the p-values using the Benjamini-Hochberg correction^49^, with results visualized in Supplementary Fig. 6. Fig. 4 highlights the ARS for traits with significant corrected p-values (*<* 0.05). Due to differences in population representation, under-represented populations such as the Middle East and America show wider confidence intervals, while more represented populations, such as the UK, exhibit narrower intervals.

Overall, there are marked differences in ARS between continental ancestries, with UK separated from Europe due to increased sample size and hence has power to make this contrast. For hay fever/allergic rhinitis, UK has significantly higher risk than that found in Europe, South Asia, and the Middle East. Psoriasis is associated with reduced risks in Africa and the Middle East, aligning with World Health Organization reports indicating lower risks in Egypt and Tanzania^50^. East Asia has the highest risk for pulmonary embolism which contrasts a significantly lower risk in the Middle East. We found significantly higher East Asia risk for high cholesterol and lower African risk for prolapsed disc/slipped disc. Other phenotypes without significant overall ARS may have indicative trends (Supplementary Fig. 5).

It should be noted that these associations are found in UK residents and are based on ancestry patterns at SNPs identified in GWAS in Europeans. They therefore may not be expected to reflect observed trait associations outside of the UK. The variability in genetic risks for these phenotypes, particularly those with significant overall ARS, remains largely underexplored due to the limited availability of genetic data across global populations.

## Discussion

In this study, we summarize the current haplotype-centric framework to understand trait associations, comparing associations of traditional PCs with haplotype-based within-sample HCs in representing genome-wide ancestry and controlling for population structure in GWAS.

We show that HCs capture finer-scale ancestral signals than those captured by PCs, thus providing a more detailed representation of genetic ancestry. HCs explain a higher proportion of variance across many phenotypes than PCs, by a factor of 2 or more for social variables such as ‘educational attainment’, ‘age of first birth’, and ‘alcohol taken with meals’. When predicting birth country, HC-based models achieve a significantly higher average recall rate than those based on PCs.

In GWAS, controlling for population structure is important to avoid false positive associations. The similar performance of HCs and PCs for many traits implies that the information needed for ancestry correction is present in PCs. These results hold for relatively unadmixed and homogeneous populations like the UK Biobank, whereas HCs may be more powerful when correcting for population structure in complex admixtures between genetically separated populations.

The orthogonal patterns of HCs^17^ indicate the nonlinear relationship of HCs would contain more useful information, especially in capturing admixtures. While this study focuses on the applications in the UK Biobank where the majority of participants are white British, applying the same methods in other biobanks with more diverse populations may show a greater advantage for HCs.

When SNPs have uncertain ancestry assignment, our results caution against the application of methods such as Tractor that try to infer differing effect sizes by population. Instead, we recommend focusing on quantities that are fully specified under uncertainty. We introduced a non-parametric hypothesis test for the significance of ARS which quantifies the genetic risk of ancestries in *admixed* datasets. We showed that ARS varies for numerous diseases and traits among continental populations, which previous studies have linked to population-specific selection^36^. This highlights the importance of population-specific medical research to build a comprehensive understanding of different diseases.

Our study also has some limitations. Although the reference panel we constructed from public-available datasets is comprehensive, it does not capture all the genetic diversity in global populations due to insufficient sample sizes. Improving reference datasets is critical to reducing the uncertainty of local ancestry inference, particularly for under-represented groups. Furthermore, our simulations highlighted the difficulty in estimating ancestry-specific effect sizes without precise local ancestry information. To address this, future work could develop scalable methods for local ancestry inference that account for admixed reference populations, or statistical models that can effectively use local ancestry probabilities to estimate ancestry-specific effects.

This study leads to two recommendations. Firstly, HCs improve genetic association with phenotypes compared to PCA, with application in GWAS and genome-wide prediction. Secondly, ARS is a useful and unbiased measure of genetic association with ancestry in admixed populations that use local ancestry and for which we now provide a robust association test. Conversely, we found that ancestry-specific effect sizes were underspecified in admixed populations with uncertain local ancestry calls. In conclusion, this study reviews the uses of haplotypes and local ancestry for understanding trait associations through a thorough examination of what works in the UK Biobank data, and what does not.

## Data Availability Statement

The UK Biobank data can be accessed by approved researchers through https://www.ukbiobank.ac.uk. We used the UK Biobank data under project 81499. POBI was accessed using accession no. EGAS00001000672. The HapMap3 variants list can be accessed at https://ftp.ncbi.nlm.nih.gov/hapmap/. All genetic data used in constructing the reference panel is provided by third parties and is available for use by others. The genetic map build GRCh37/hg19 is available from https://bochet.gcc.biostat. washington.edu/beagle/genetic maps/. The UK and world map data are available at https://gadm.org and through R package ‘rworldmap’ at https://cran.r-project.org/web/packages/rworldmap/index.html.

## Code Availability

The code for the simulation of ancestry-specific GWAS is available on GitHub at https://github.com/ YaolingYang/sim ancestry GWAS. The pipeline of using SparsePainter to paint UK Biobank individuals and using PBWTpaint to compute HCs are available on GitHub at https://github.com/YaolingYang/SparsePainter/tree/main/painting-pipeline.

## Acknowledgments

We thank the participants in the UK Biobank (UKB), HAPMAP3, HGDP, POBI, Busby Europe, Pagani Africa, Peterson Africa and Schlebusch Africa. Y.Y. was supported by China Scholarship Council [grant number 202108060092]. This work was carried out using the computational facilities of the Advanced Computing Research Centre, University of Bristol - http://www.bris.ac.uk/acrc.

## Author Contribution Statement

Y.Y. and D.J.L. conceived and designed the project and methodology. D.J.L. supervised the project.

Y.Y. and D.J.L. developed the methodology. Y.Y. designed the pipeline for painting the UK Biobank and performed simulations and data analysis under the supervision of D.J.L. Y.Y. wrote the initial manuscript draft. Y.Y. and D.J.L. wrote, reviewed, discussed and revised the subsequent versions of the manuscript, and agreed with the submitted manuscript.

## Ethical Approval

This study used data from the UK Biobank (application number 80499), HAPMAP3, HGDP, POBI, Busby Europe, Pagani Africa, Peterson Africa and Schlebusch Africa, which adhered to ethical standards and guidelines established for human genome research. The UK Biobank has obtained ethical approval from the North West Multi-centre Research Ethics Committee. The other datasets provide publicly available genetic data that do not require additional institutional review board approval for secondary analysis. The study was designed and conducted in compliance with all the relevant regulations regarding the use of human study participants and the criteria set by the Declaration of Helsinki.

