## Supplementary Information for "From individuals to ancestries: towards attributing trait variation to haplotypes"

### Supplementary Note

#### Converting educational attainment from categorical variable to continuous variable

Educational attainment is the only categorical phenotype included in our analysis, we convert them into continuous scores using the International Standard Classification for Education (ISCED) definition[1,2] below:

$$ES = \begin{cases} 7, \text{ if ''none of the above''} \\ 10, \text{ if ''CSEs or equivalent'' or ''O levels/GCSEs or equivalent''} \\ 13, \text{'' A levels/AS levels or equivalent''} \\ 15, \text{ Other professional qualifications eg: nursing, teaching} \\ 19, NVQ \text{ or HND or HNC or equivalent} \\ 20, \text{ College or University degree} \end{cases}$$

### Supplementary Table Legends

**Supplementary Table 1:** Distribution of n=20,092 individuals by birthplace and self-reported ethnic group in the study assessing the performance of HCs and PCs to predict birthplaces from the UK Biobank.

**Supplementary Table 2:** 93 world-wide ancestries and their abbreviations as summarized from public-available reference datasets.

**Supplementary Table 3:** Average out-of-sample $R^2$ explained by the top 150 HCs and top 18 PCs and the optimal number of top HCs and PCs for continuous phenotypes.

**Supplementary Table 4:** Average out-of-sample Mcfadden's $R^2$ explained by the top 150 HCs and top 18 PCs and the optimal number of top HCs and PCs for binary phenotypes.

### Supplementary Figures

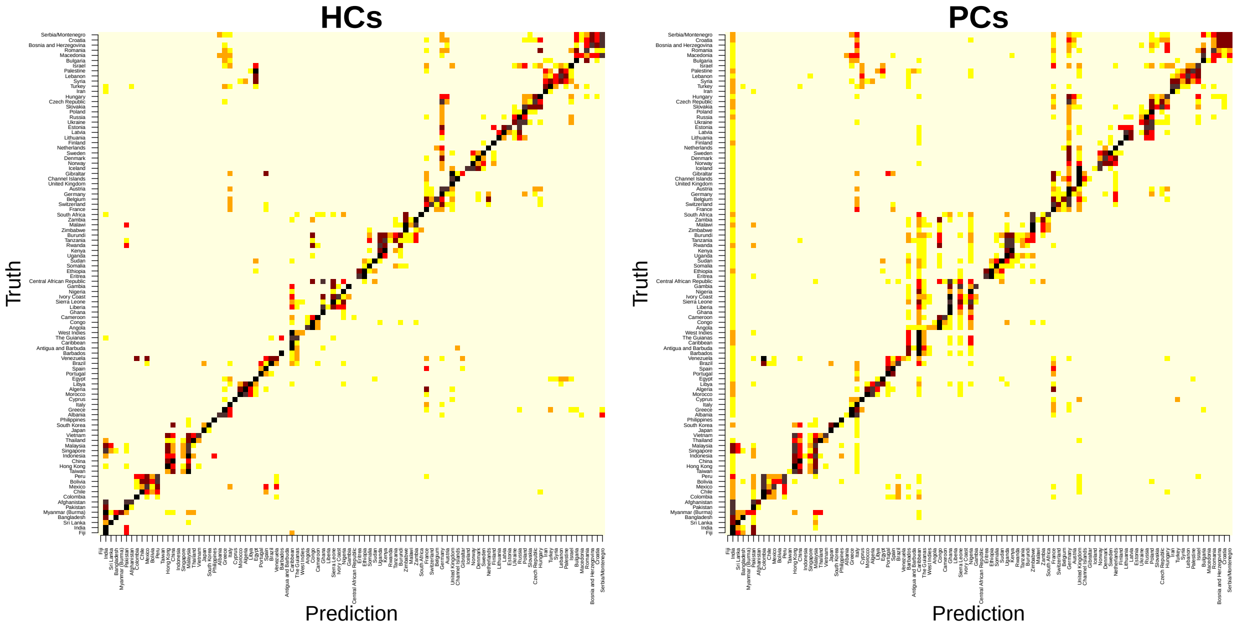


**Supplementary Fig. 1**: **Confusion matrices for the performance of HCs and PCs on predicting worldwide birthplaces visualized in heatmaps.**

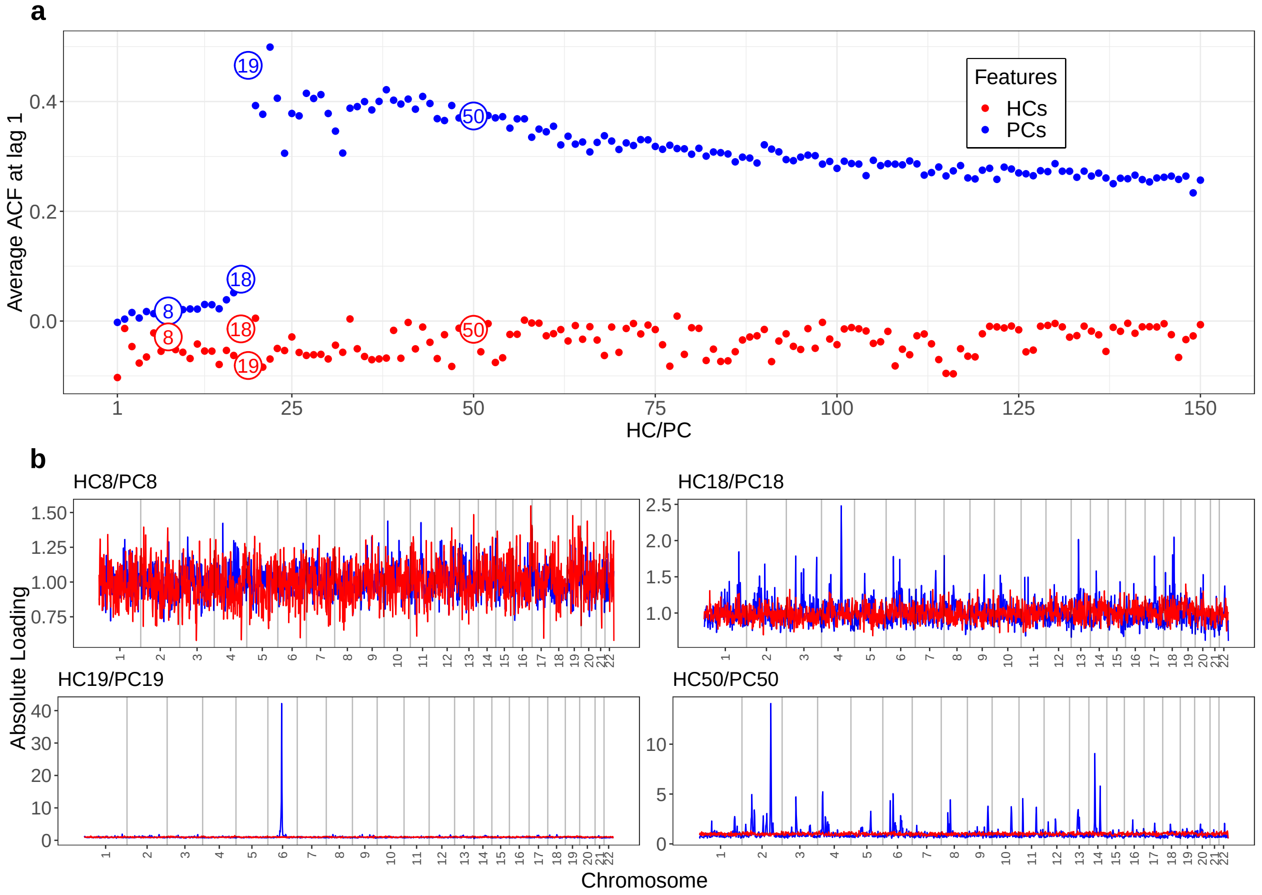


**Supplementary Fig. 2**: **LD structure as a function of HCs/PCs in the UK Biobank.** a, average auto-correlation function (ACF) at lag 1 of the absolute value of loadings for the top 150 HCs and PCs. b, aggregated absolute value of loadings from highlighted HCs/PCs (red/blue in a) throughout the genome with SNPs aggregated into bins of 100. A total of n=406,773 individuals from the UK Biobank are included in this analysis.

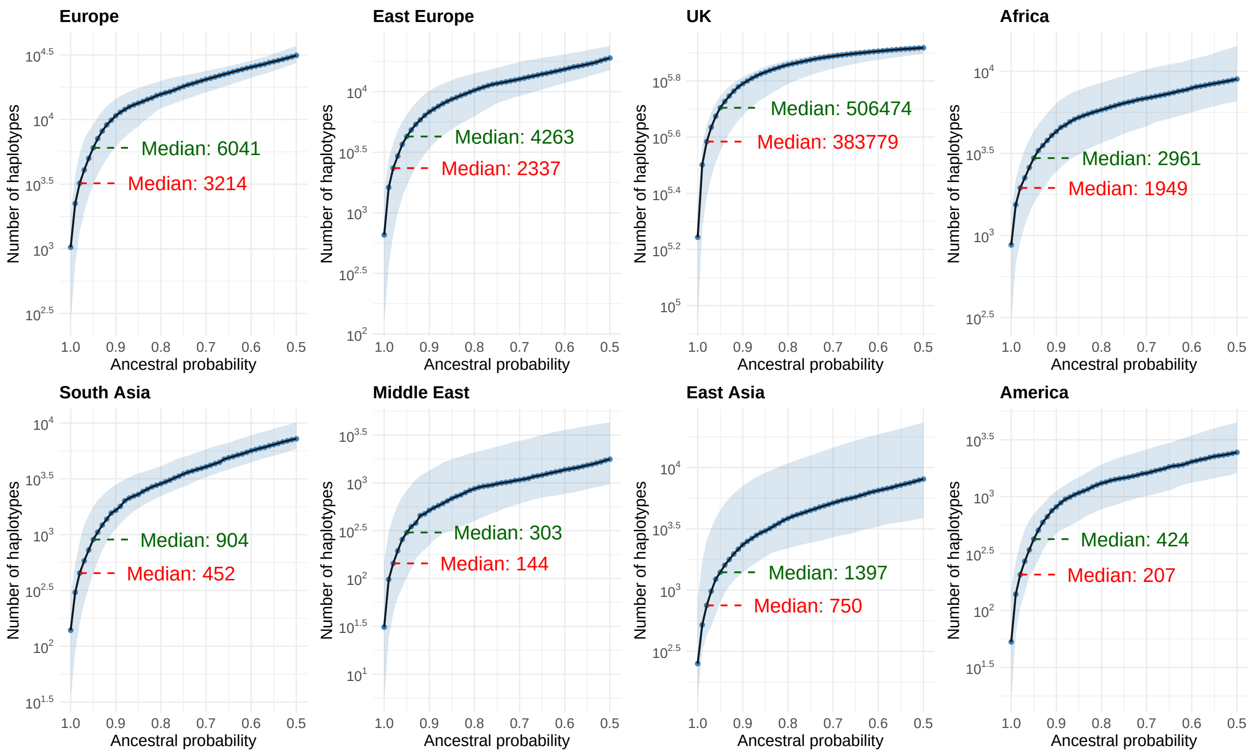


**Supplementary Fig. 3**: **Distribution of painted UK Biobank samples across ancestral probabilities by continent.** The x-axis represents the local ancestry probabilities, and the y-axis represents the number of haplotypes measured as two times the number of individuals painted in the UK Biobank on a log10 scale. There are a total of n=925,388 haplotypes. Each plot displays the median cumulative haplotype count (blue dashed line) along with the corresponding 95% confidence intervals (shaded regions) at varying local ancestry probability thresholds, based on all pruned significant SNPs across 35 phenotypes (investigated in the ARS analysis) for 8 ancestries. Median values at the 98% and 95% probability thresholds are annotated.

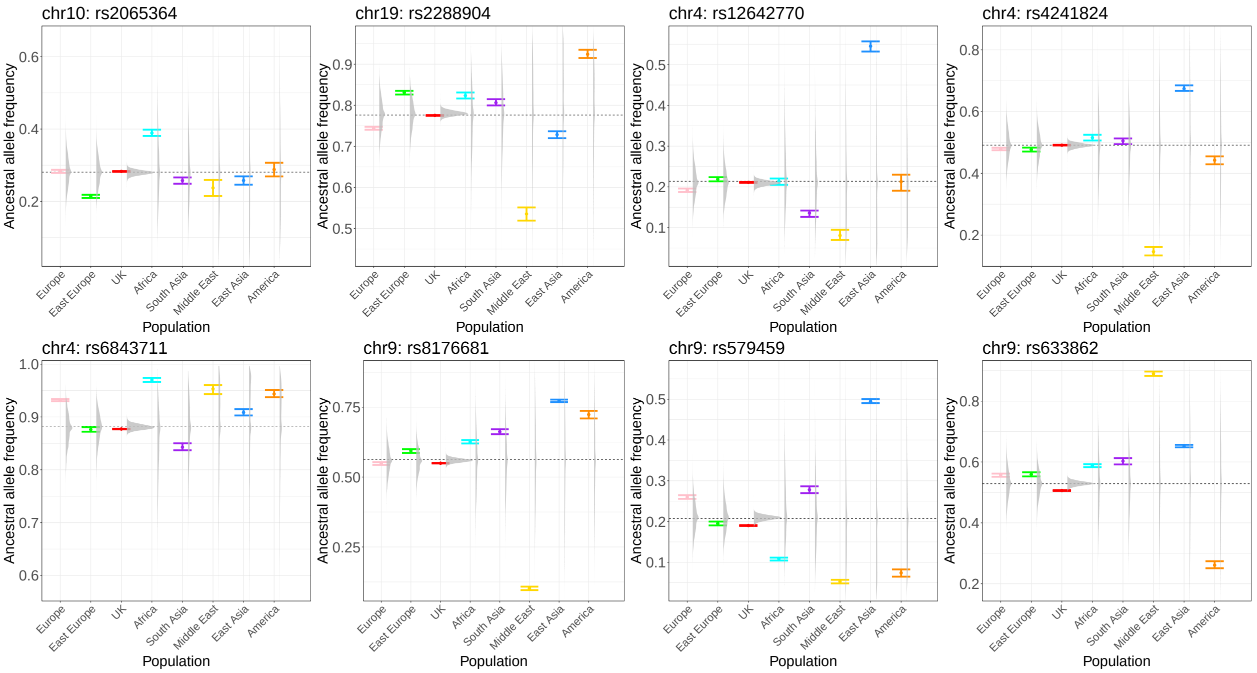


**Supplementary Fig. 4**: **Simulated distribution of ancestral allele frequency for pulmonary embolism.** The error bars represent the 95% confidence interval of ancestral allele frequency. The distribution of simulated allele frequency for each population of all the matched SNPs in the UK Biobank is shown as a raincloud plot. The dashed black lines represent the genome-wide allele frequency.

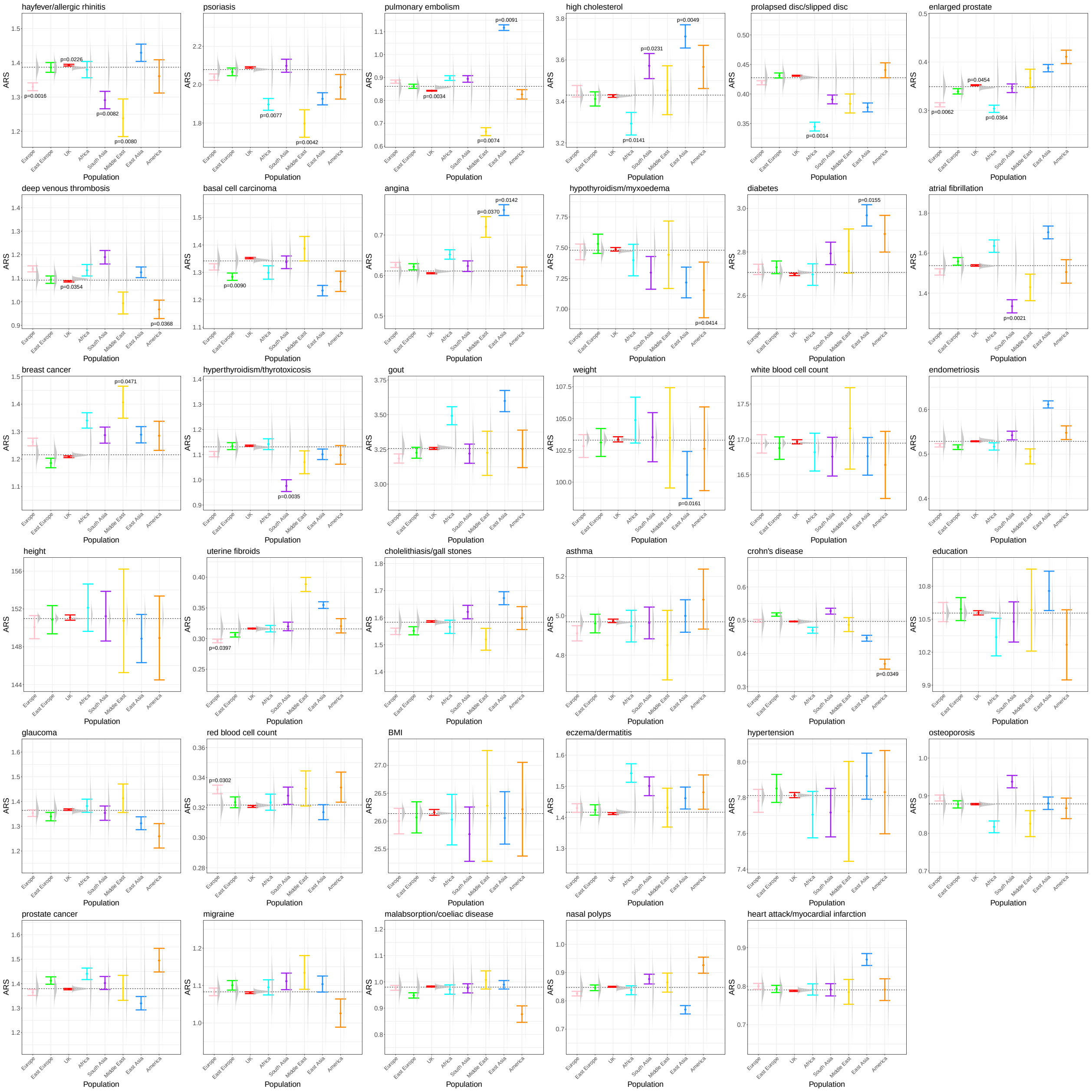


**Supplementary Fig. 5**: **ARS for 35 phenotypes ordered by the overall significance level.** ARS are computed from n=462,694 individuals in the UK Biobank. The error bars represent the 95% confidence interval of ARS. The distribution of simulated ARS for each population is shown as a raincloud plot. The dashed black lines represent the average ARS of all populations weighted by the population sizes. The ARS of populations with raw p-values significant at 5% computed from the two-sided empirical test are annotated.

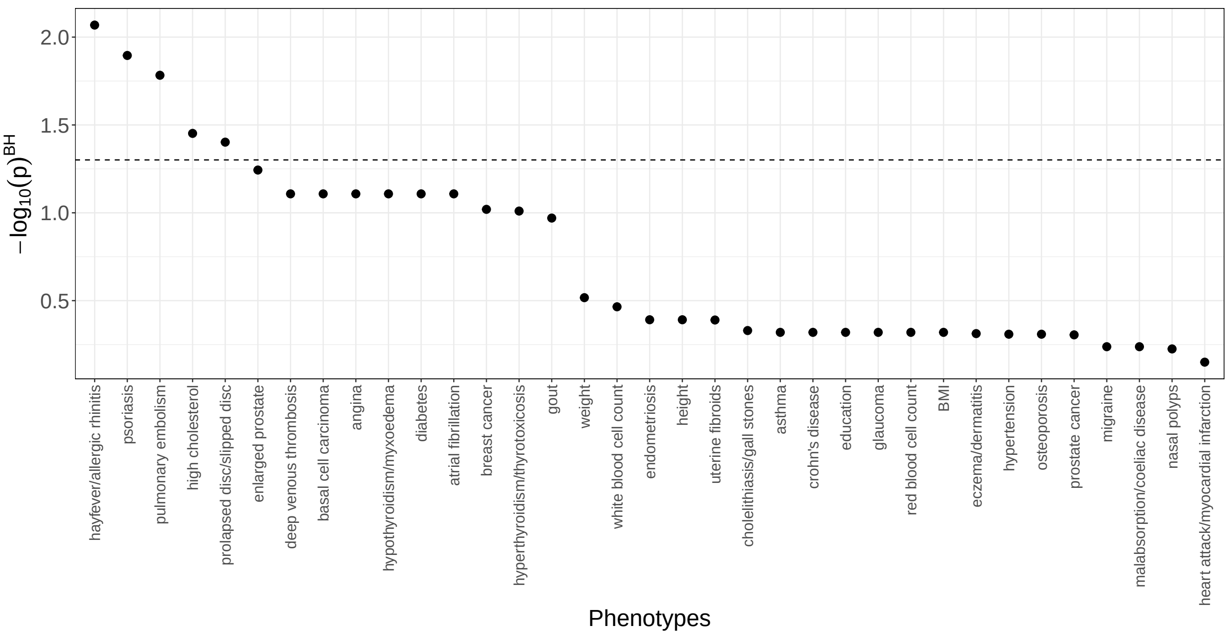


**Supplementary Fig. 6**: **Phenotype-wise association of ARS across populations using Fisher's method.** The y-axis shows the -log10 scale of the p-values derived from the ARS calculated across different populations and then aggregated using Fisher's method, i.e. chi-squared test (see Methods).

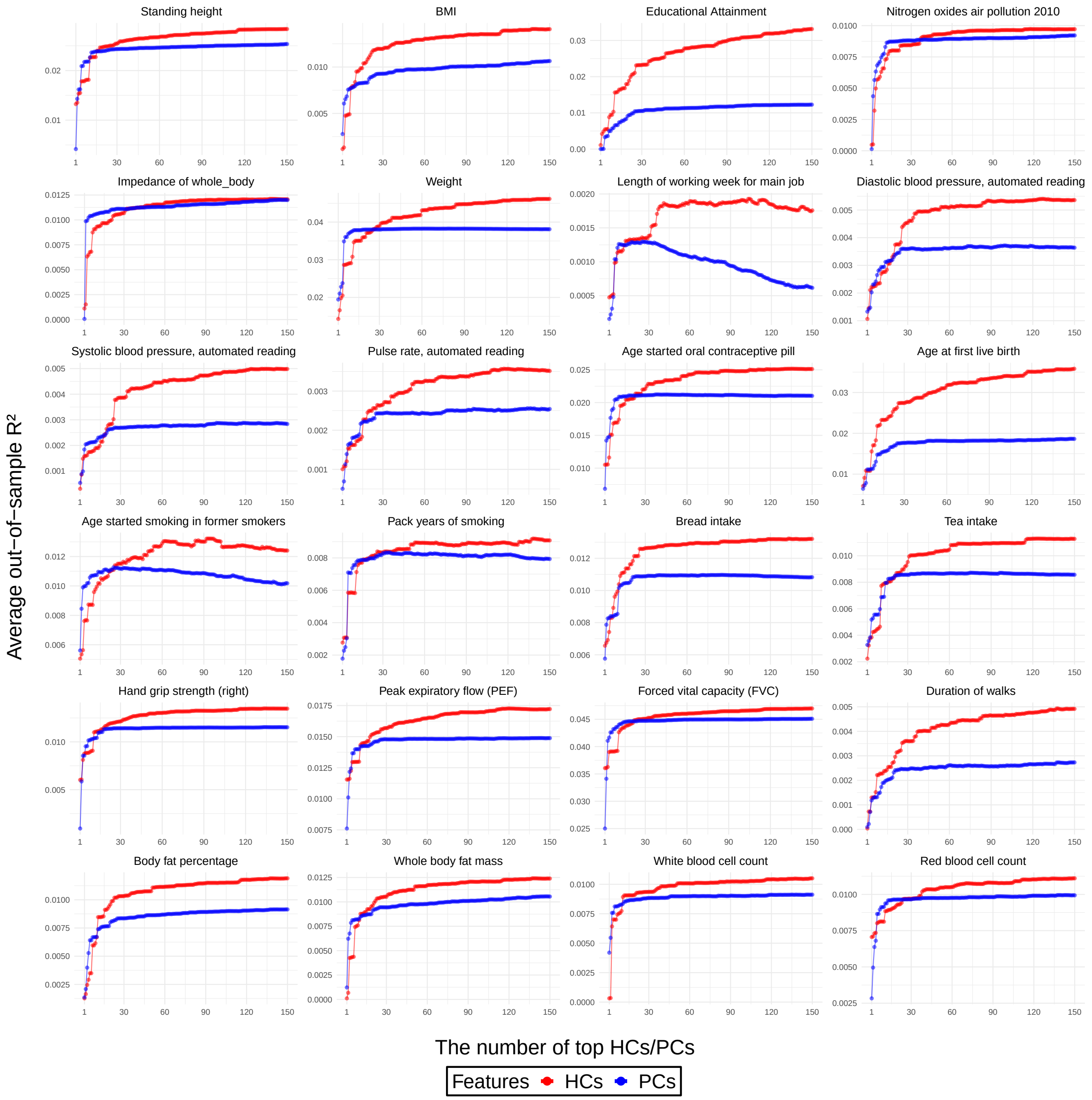


**Supplementary Fig. 7**: **Average out-of-sample $R^2$ explained by different numbers of top HC-s/PCs for 24 continuous phenotypes.** This study includes n=406,773 UK Biobank individuals.

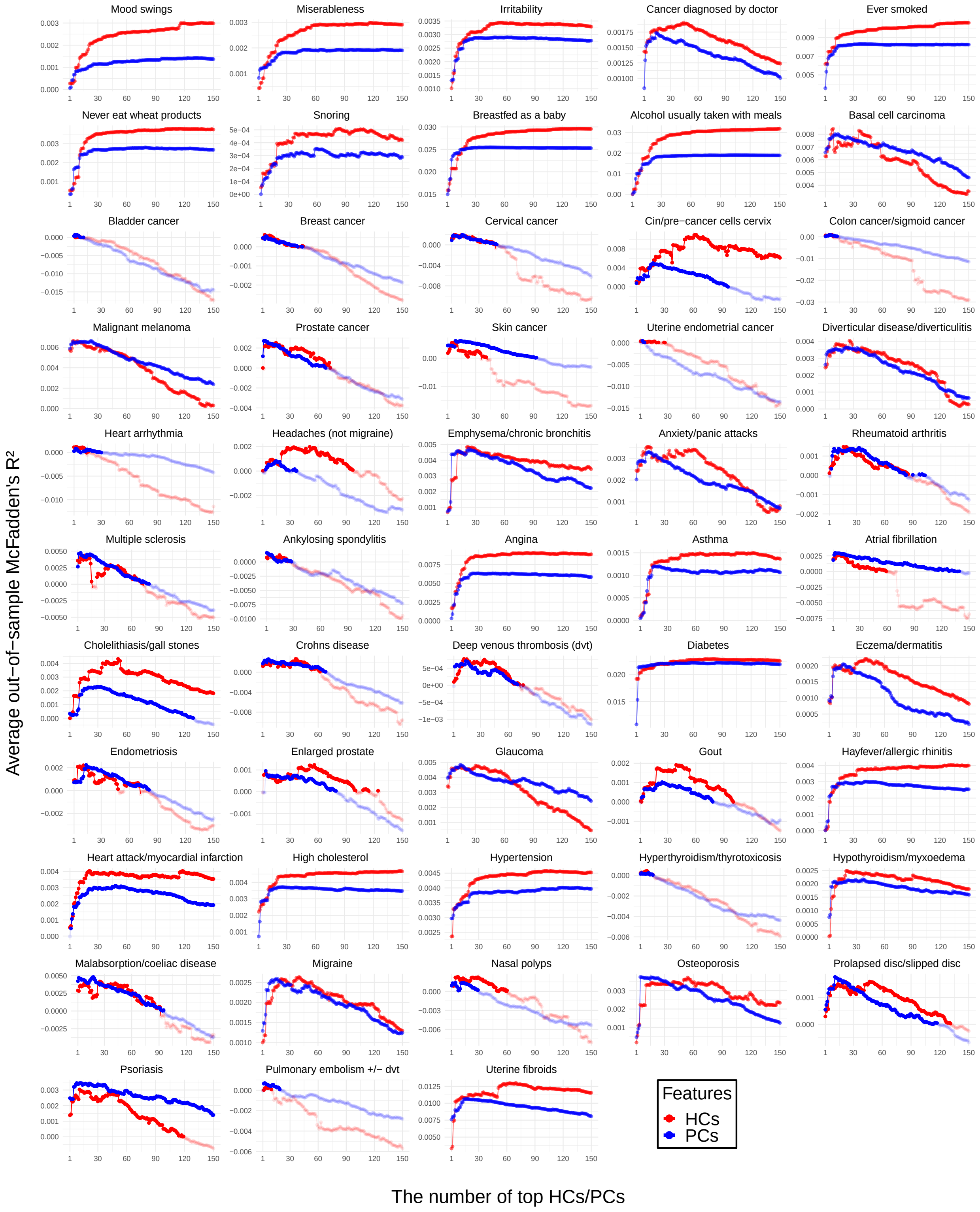


**Supplementary Fig. 8**: **Average out-of-sample $R^2$ explained by different numbers of top HCs/PCs for 53 binary phenotypes.** The points and lines are faded for negative average out-of-sample $R^2$. This study includes n=406,773 UK Biobank individuals.

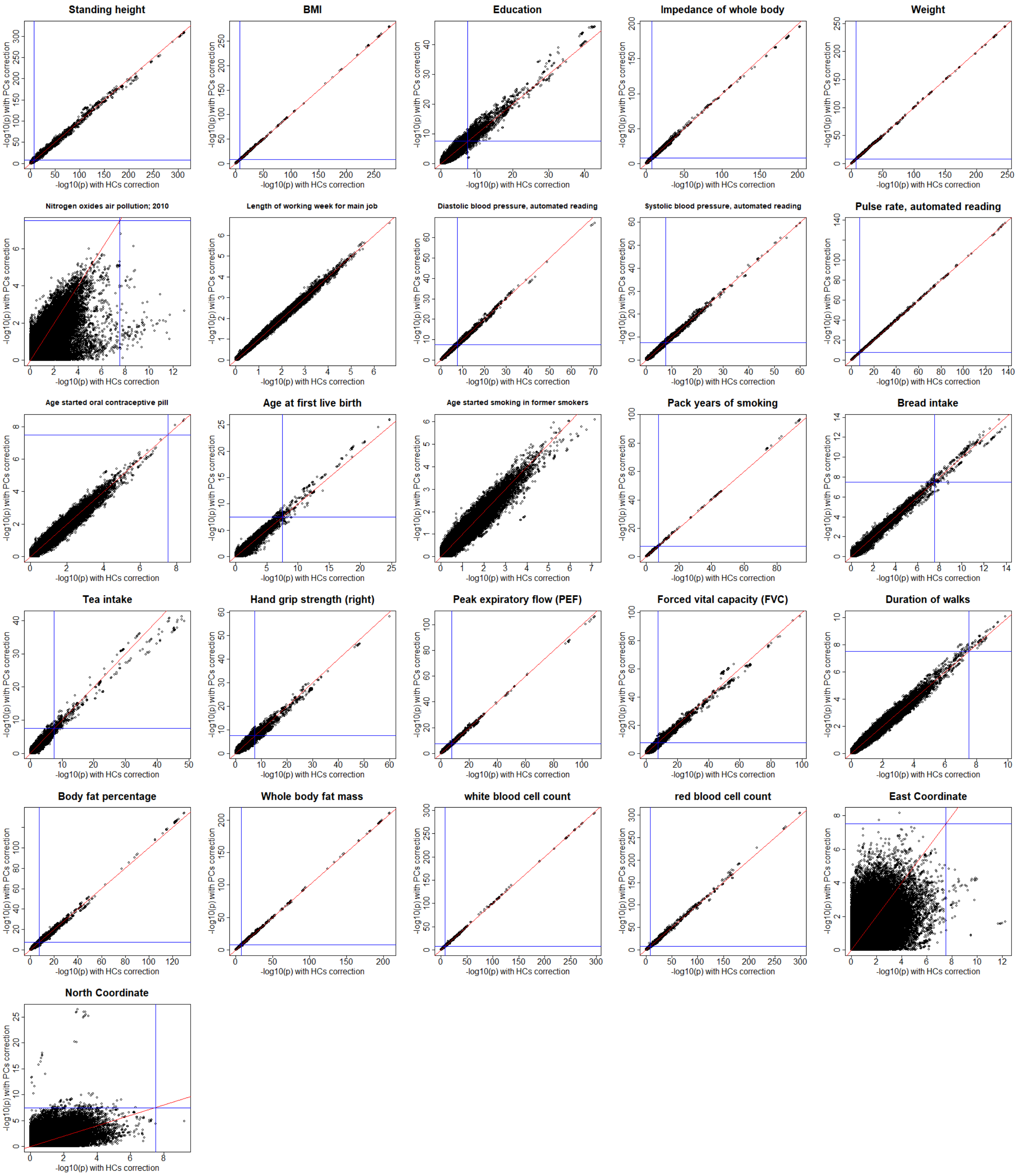


**Supplementary Fig. 9**: **Comparison between HC-corrected and PC-corrected GWAS for 26 continuous phenotypes.** Each point represents its -log10(P-value) from GWAS corrected by the top 18 HCs or PCs. This study includes n=406,773 UK Biobank individuals.

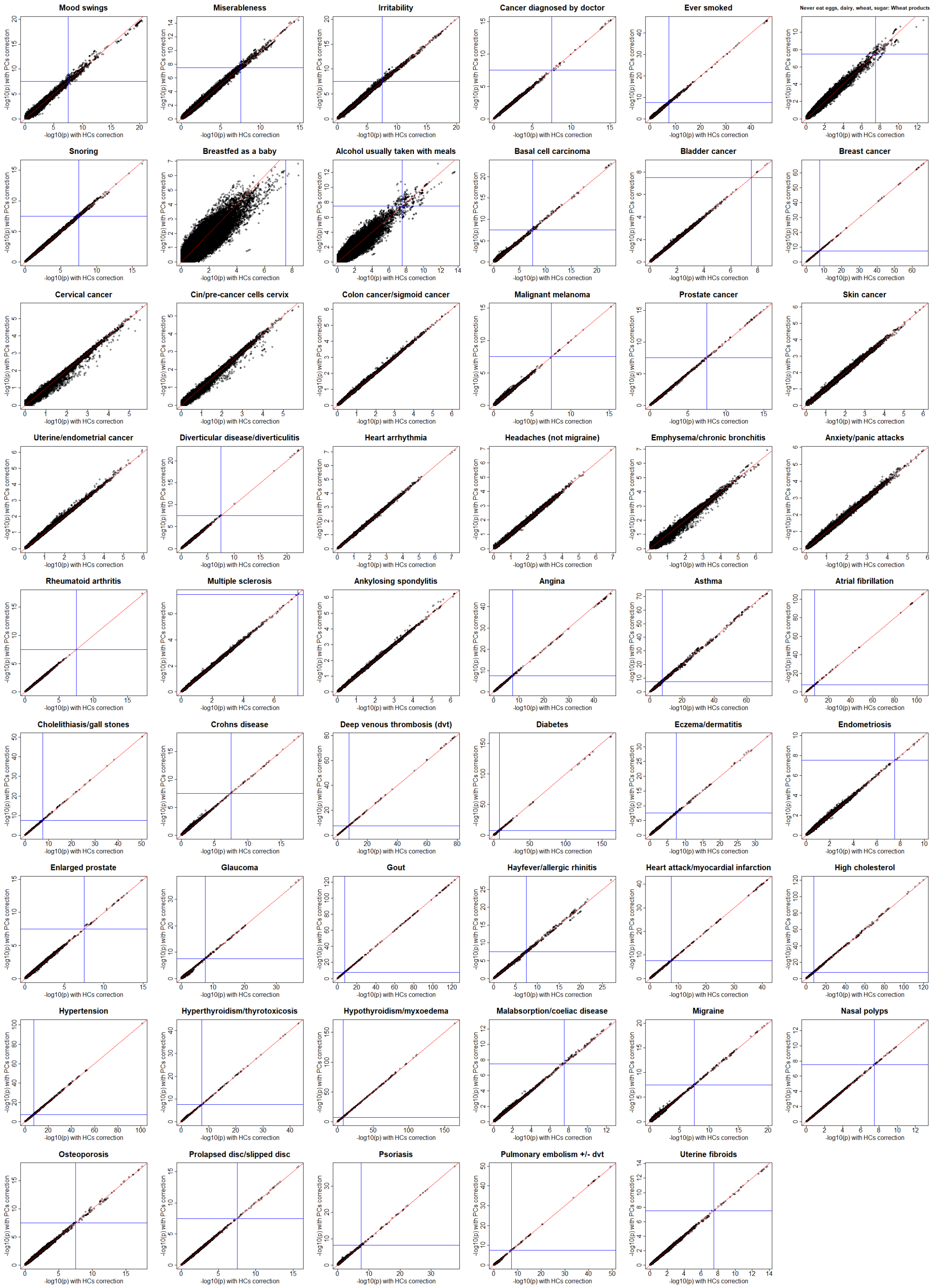


**Supplementary Fig. 10**: **Comparison between HC-corrected and PC-corrected GWAS for 53 binary phenotypes.** Each point represents its -log10(P-value) from GWAS corrected by the top 18 HCs or PCs. This study includes n=406,773 UK Biobank individuals.

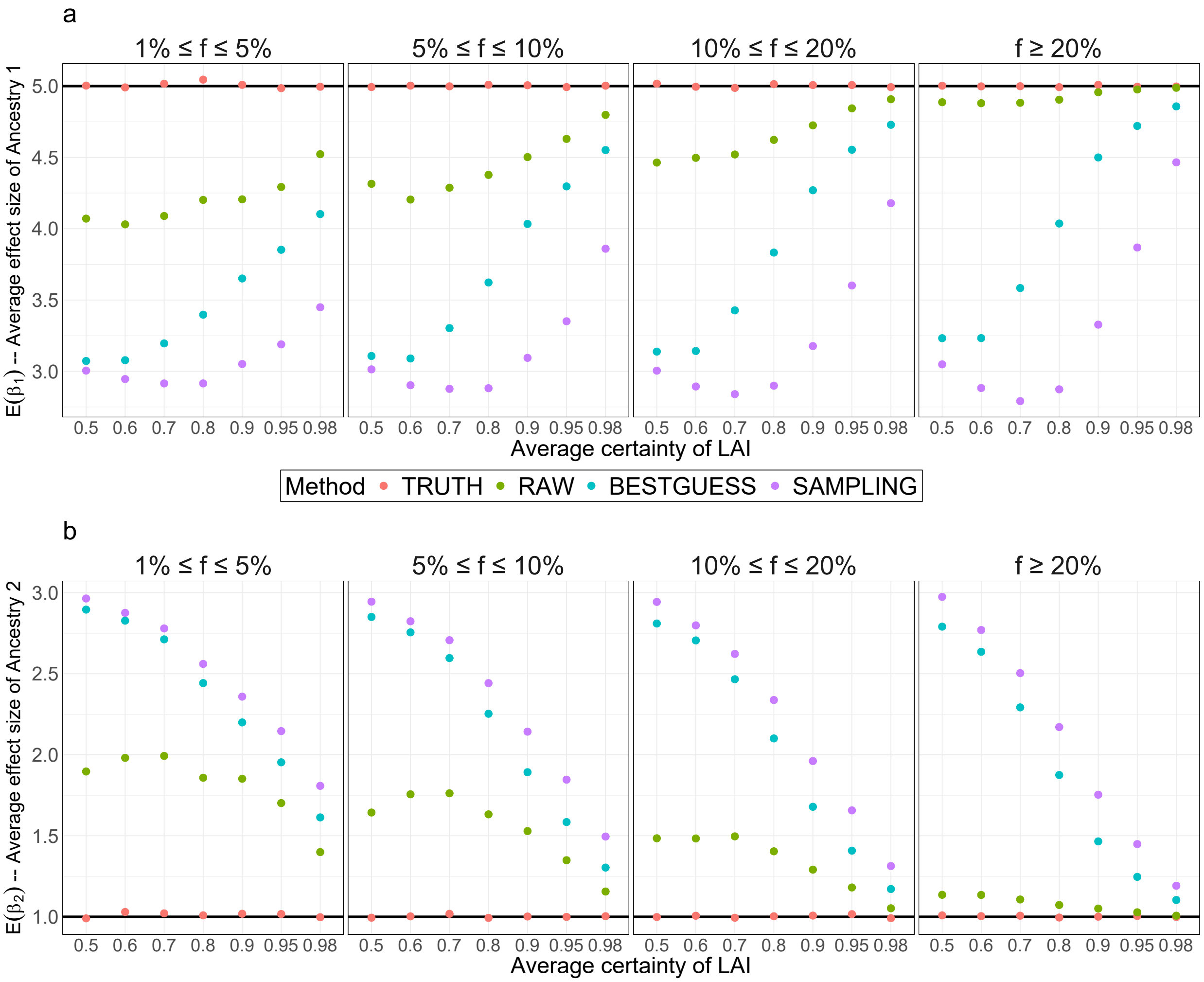


**Supplementary Fig. 11**: **Comparison of the average estimated effect size of ancestries from the Tractor model with different methods to represent local ancestries under $\mathbf{E(A^*_{i1d}) = 0.25}$.** $E(A^*_{i1d})$ denotes the average probability of Ancestry 1. The x-axis represents the average certainty of LAI (see Methods) and the y-axis represents the average estimated effect sizes for Ancestry 1 (plot a) and Ancestry 2 (plot b). Different MAF thresholds $f$ are compared: $1\% \leq f \leq 5\%$, $5\% \leq f \leq 10\%$, $10\% \leq f \leq 20\%$, and $f \geq 20\%$. The simulation was repeated 1,000 times with n=20,000 diploid individuals.

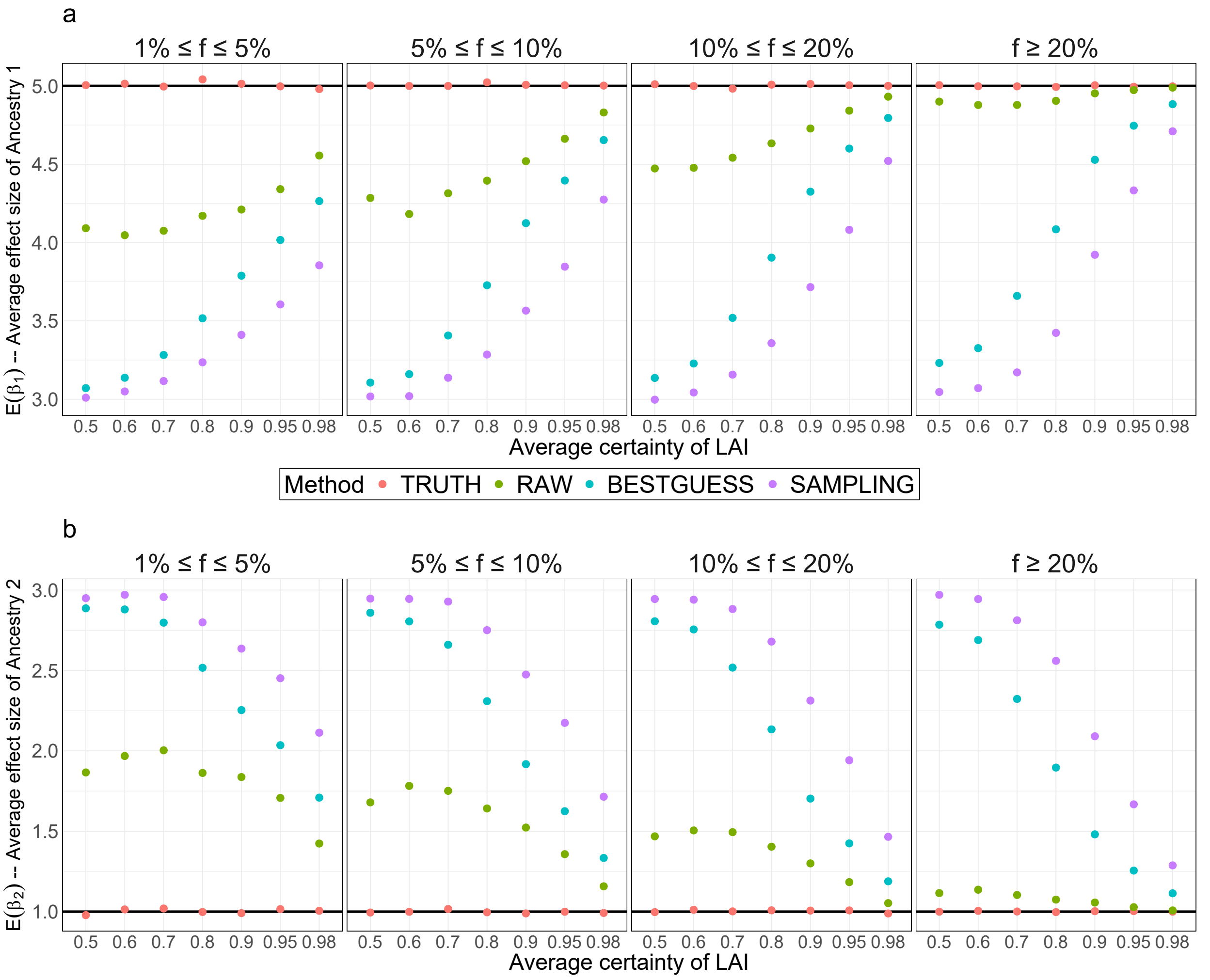


**Supplementary Fig. 12**: **Comparison of the average estimated effect size of ancestries from the Tractor model with different methods to represent local ancestries under $\mathbf{E(A^*_{i1d}) = 0.5}$.** $E(A^*_{i1d})$ denotes the average probability of Ancestry 1. The x-axis represents the average certainty of LAI (see Methods) and the y-axis represents the average estimated effect sizes for Ancestry 1 (plot a) and Ancestry 2 (plot b). Different MAF thresholds $f$ are compared: $1\% \leq f \leq 5\%$, $5\% \leq f \leq 10\%$, $10\% \leq f \leq 20\%$, and $f \geq 20\%$. The simulation was repeated 1,000 times with n=20,000 diploid individuals.

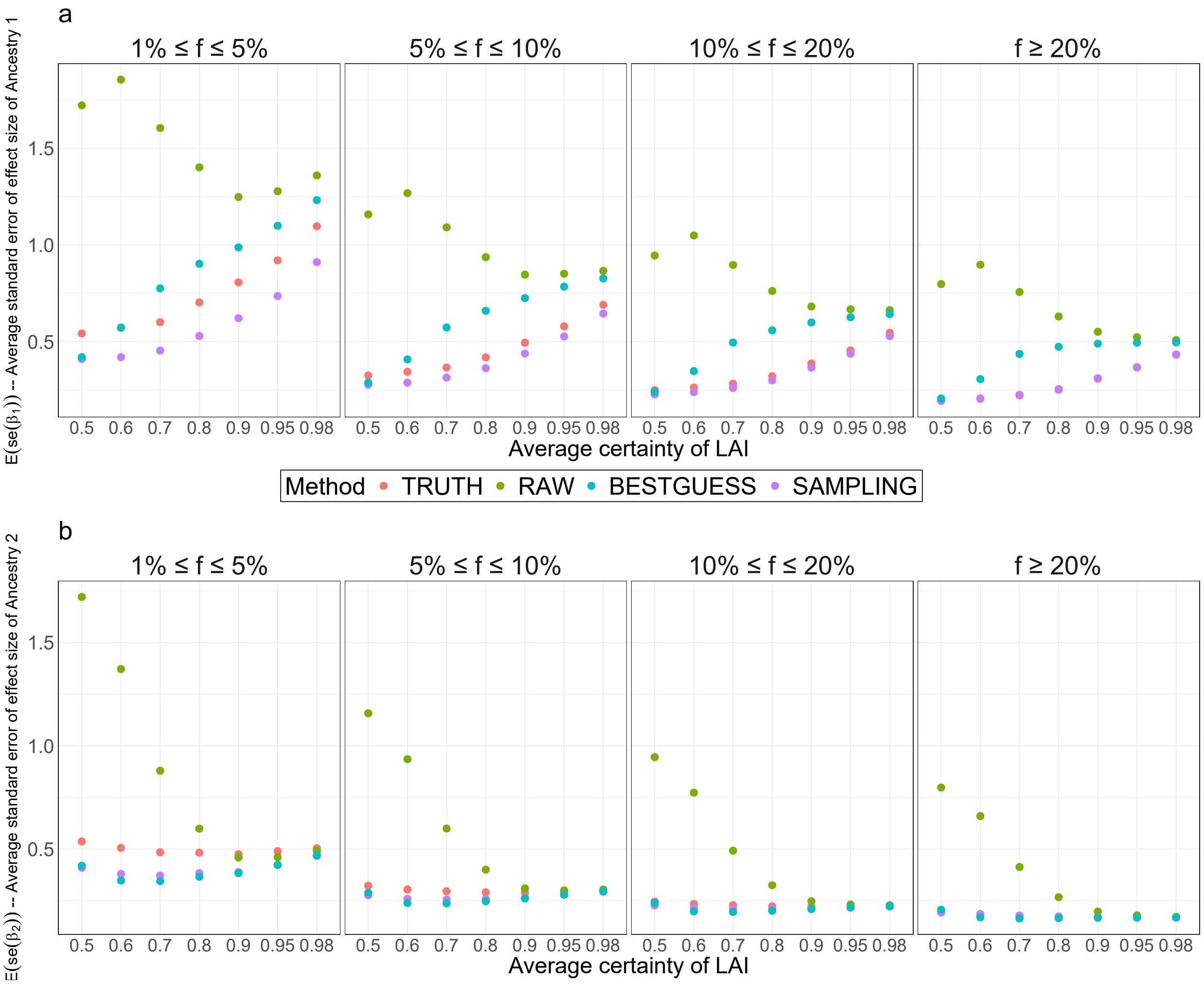


**Supplementary Fig. 13**: **Comparison of the standard error of the estimated effect size of ancestries from the Tractor model with different methods to represent local ancestries under $\mathbf{E(A^*_{i1d}) = 0.1}$.** $E(A^*_{i1d})$ denotes the average probability of Ancestry 1. The x-axis represents the average certainty of LAI (see Methods) and the y-axis represents the average standard error of the estimated effect size of Ancestry 1 (plot a) and Ancestry 2 (plot b). Different MAF thresholds $f$ are compared: $1\% \le f \le 5\%$, $5\% \le f \le 10\%$, $10\% \le f \le 20\%$, and $f \ge 20\%$. The simulation was repeated 1,000 times with n=20,000 diploid individuals.

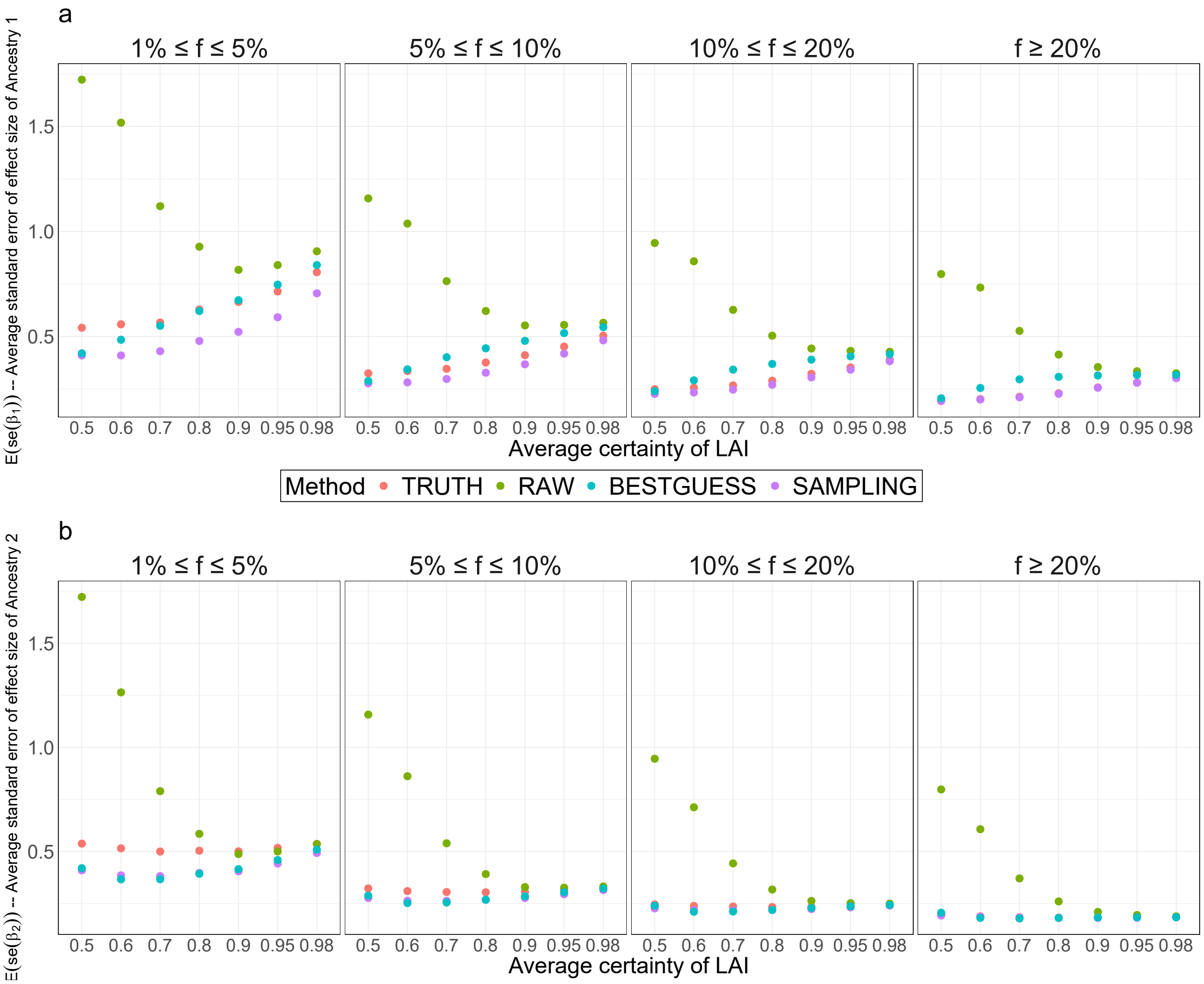


**Supplementary Fig. 14**: **Comparison of the standard error of the estimated effect size of ancestries from the Tractor model with different methods to represent local ancestries under $\mathbf{E(A^*_{i1d}) = 0.25}$.** $E(A^*_{i1d})$ denotes the average probability of Ancestry 1. The x-axis represents the average certainty of LAI (see Methods) and the y-axis represents the average standard error of the estimated effect size of Ancestry 1 (plot a) and Ancestry 2 (plot b). Different MAF thresholds $f$ are compared: $1\% \leq f \leq 5\%$, $5\% \leq f \leq 10\%$, $10\% \leq f \leq 20\%$, and $f \geq 20\%$. The simulation was repeated 1,000 times with n=20,000 diploid individuals.

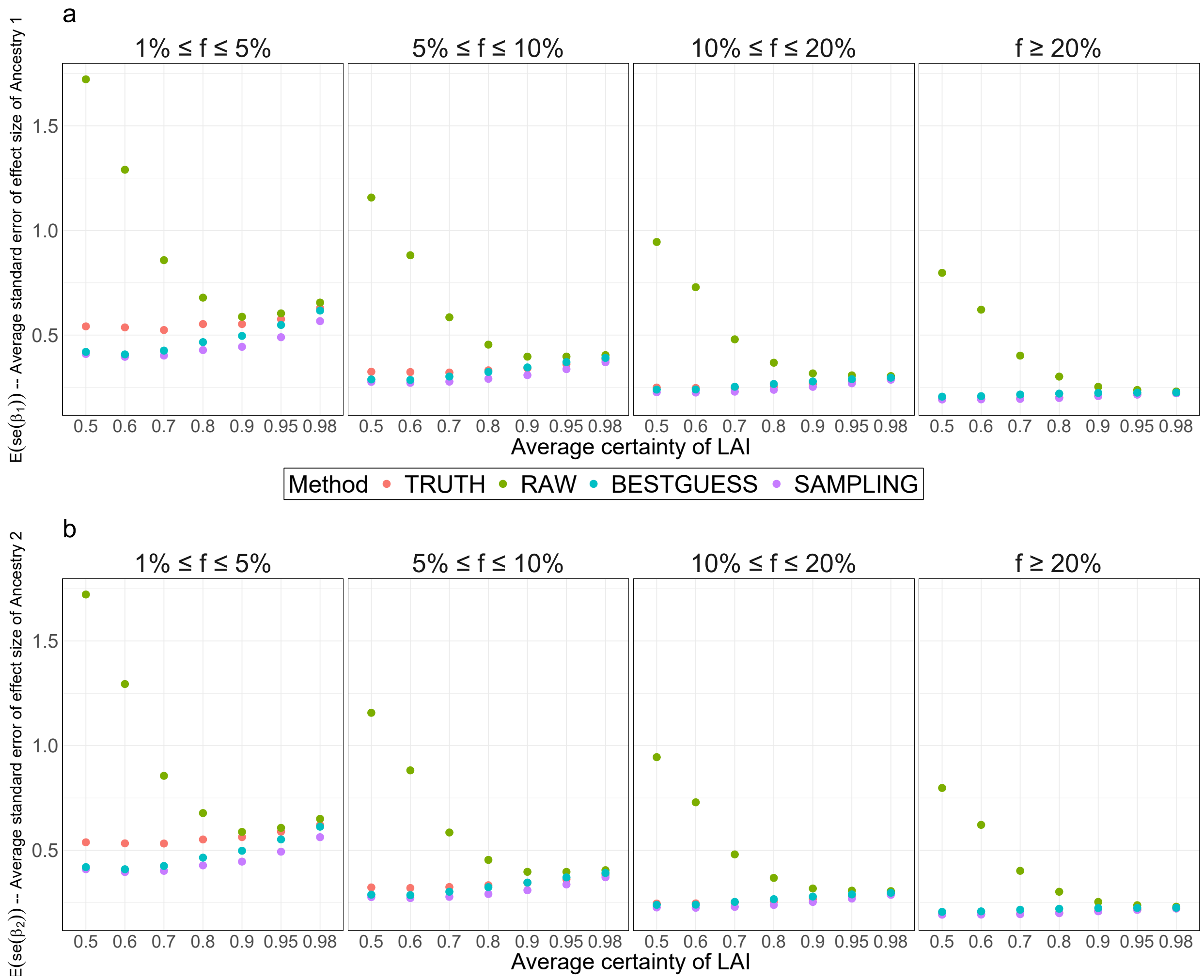


**Supplementary Fig. 15**: **Comparison of the standard error of the estimated effect size of ancestries from the Tractor model with different methods to represent local ancestries under $\mathbf{E(A^*_{i1d}) = 0.5}$.** $E(A^*_{i1d})$ denotes the average probability of Ancestry 1. The x-axis represents the average certainty of LAI (see Methods) and the y-axis represents the average standard error of the estimated effect size of Ancestry 1 (plot a) and Ancestry 2 (plot b). Different MAF thresholds $f$ are compared: $1\% \leq f \leq 5\%$, $5\% \leq f \leq 10\%$, $10\% \leq f \leq 20\%$, and $f \geq 20\%$. The simulation was repeated 1,000 times with n=20,000 diploid individuals.
